# Plasma Proteomic Networks Reveal Shared Biology with Brain Linked to Alzheimer’s Disease Pathology

**DOI:** 10.64898/2026.05.26.26353866

**Authors:** Qi Guo, Lingyan Ping, Saima Rathore, Duc M. Duong, Anantharaman Shantaraman, Edward J. Fox, Erik C.B. Johnson, James J. Lah, Allan I. Levey, Nicholas T. Seyfried

**Affiliations:** Department of Biochemistry, School of Medicine, Emory University, Atlanta, GA 30322, USA; Center for Neurodegenerative Disease, School of Medicine, Emory University, Atlanta, GA 30322, USA; Goizueta Alzheimer’s Disease Research Center, School of Medicine, Emory University, Atlanta, GA 30322, USA; Department of Neurology, School of Medicine, Emory University, Atlanta, GA 30322, USA; Department of Biomedical Informatics, School of Medicine, Emory University, Atlanta, GA 30322, USA

**Keywords:** Alzheimer’s disease, amyloid, biomarkers, cerebrospinal fluid, plasma, phosphorylated tau, proteomics, microbead enrichment, data-independent acquisition (DIA), mass spectrometry

## Abstract

Alzheimer’s disease (AD) drives widespread molecular changes beyond the brain that are increasingly detectable in plasma. To map plasma proteomic signatures of AD in a broadly unbiased manner with high depth and reproducibility, we profiled plasma from 214 individuals spanning cognitively normal controls, mild cognitive impairment, and AD using microbead-based enrichment and data-independent acquisition mass spectrometry (DIA-MS). We reliably quantified 5,823 proteins across samples, and network analysis identified 29 plasma modules enriched for functions related to lipid metabolism, extracellular matrix remodeling, immune signaling, mitochondrial function, and proteostasis. Several modules were associated with cognition, APOE4, sex, race, and cerebrospinal fluid (CSF) amyloid and tau biomarkers. Among 129 individuals with paired CSF and plasma biomarker measurements, over 1,500 proteins differed between CSF biomarker–positive and –negative groups, including amyloid-linked matrisome proteins such as SMOC1, FRZB, SPON1 and CTHRC1. A 10-protein plasma panel classified CSF biomarker positivity with performance similar to plasma pTau217 (AUC = 0.91), and combining both improved accuracy (AUC = 0.99). Integration with a human brain proteomic network revealed that two-thirds of plasma modules were preserved in brain, with many AD-altered modules changing concordantly across compartments. This study establishes a scalable DIA-MS plasma proteomics platform that captures systemic and brain-linked AD biology and identifies complementary biomarkers beyond phosphorylated tau.

## Introduction

Alzheimer’s disease (AD) is characterized by extracellular amyloid-β (Aβ) deposition, intracellular accumulation of hyperphosphorylated tau (pTau), and progressive neurodegeneration leading to cognitive decline (1–3). Although cerebrospinal fluid (CSF) and positron emission tomography (PET) biomarkers remain the gold standard for detecting Aβ and tau pathology (4, 5), their costs, invasiveness, and limited availability have accelerated the search for scalable blood-based biomarkers (6, 7). Phosphorylated tau species (pTau) at specific residues, especially pTau at threonine 217 in plasma (pTau217), correlate strongly with plaque pathology in the brain, whereas proteins like neurofilaments and glial fibrillary acidic protein (GFAP), among others, predict progression to dementia (8–11). Yet these targeted markers represent only a narrow fraction of the biology accompanying AD, and they do not fully reflect the diverse pathways implicated in disease, including inflammation, lipid metabolism, mitochondrial dysfunction, extracellular matrix remodeling, and vascular damage (12–15).

Large-scale affinity-based proteomic platforms using either antibodies or aptamers, such as Olink and SomaScan, have expanded the landscape for plasma biomarkers by enabling high-throughput quantification of thousands of proteins from small sample volumes (16–18). These technologies have been applied in population-scale studies, including the UK Biobank and the Global Neurodegeneration Proteomics Consortium (GNPC), to identify immune, vascular, and metabolic signatures associated with cognition and dementia (19–21). However, affinity-based methods are inherently constrained by assay specificity, epitope accessibility, and nonlinear behavior across the 12–13 orders of dynamic range in plasma protein abundance (22, 23). Furthermore, they are not truly unbiased and rarely resolve proteoforms, including alternatively spliced isoforms and post-translationally modified peptides (24, 25).

Mass spectrometry (MS)-based plasma proteomics offers these advantages but has historically struggled to achieve deep proteome coverage due to the overwhelming dominance of albumin, immunoglobulins, and other high-abundance proteins (26–28). Early MS proteomic workflows attempted to address this challenge using albumin and immunoglobulin depletion, ultracentrifugation, or organic precipitation (26, 29–31). This includes our own prior heparin-enriched MS study, which quantified over 2,000 heparin-binding and ECM-related proteins in AD plasma (29). While valuable, these approaches lack the depth and scalability of affinity-based platforms, limiting their utility for large-scale network biology and biomarker discovery (27, 32, 33).

Recent technological advances now enable a fundamentally different strategy. Particle-based enrichment approaches, such as Seer’s multiplexed nanoparticles and functionalized microbeads from PreOmics, capture diverse subsets of the plasma proteome through physicochemical interactions (34–36). Unlike MS methods coupled with immunodepletion or affinity-based methods, these approaches do not remove abundant proteins or introduce antibody/aptamer biases (34–36). Instead, they redistribute the plasma proteome by selectively enriching low- and medium-abundance proteins, thereby increasing the effective dynamic range. When coupled with data-independent acquisition (DIA) on highly sensitive MS platforms, these enrichment approaches dramatically enhance proteome depth, enabling scalable discovery workflows that quantify 4,000–7,000 proteins per sample with excellent reproducibility (28, 37, 38).

In this study, we used a one-step ENRICHplus microbead-based enrichment workflow developed by PreOmics combined with relatively high-throughput DIA-MS on the Orbitrap Astral (40 samples/day) to deeply profile the plasma proteome of 214 individuals, including cognitively normal controls, MCI, and AD cases. This platform quantified 6,819 total proteins (65,045 unique peptides), with 5,823 reliably measured across samples (median coefficient of variation ∼13%). The protein measures spanned a wide dynamic range and enabled construction of a robust co-expression network comprising 29 biological modules enriched for lipid metabolism, extracellular matrix remodeling, mitochondrial function, and proteostasis. Importantly, this deep, reproducible microbead-based approach provided plasma proteome coverage exceeding that of prior depletion-based or heparin-based MS methods (29, 31, 32). It also provides both shared and complementary protein profiles relative to affinity-based platforms, capturing thousands of intracellular, vesicular, and mitochondrial proteins that are typically not detected in plasma. We further show that plasma proteomic networks capture AD-related biology associated with cognitive performance, APOE4 gene dose, demographic factors, and CSF amyloid and tau biomarkers. These plasma signatures also closely mirror brain proteomic networks across major biological pathways. Using machine learning, we identified a ten-protein plasma panel that, when combined with plasma pTau217, significantly improved classification of individuals positive for CSF AD biomarkers, highlighting the potential of targeted multi-protein assays for scalable clinical implementation. Collectively, these findings establish microbead-enriched, DIA-MS plasma proteomics as a scalable platform that overcomes longstanding limitations of MS and enables the identification of complementary plasma biomarkers beyond pTau217.

## Methods

### Plasma Samples

All participants providing plasma samples gave informed consent under protocols approved by the Institutional Review Board at Emory University. Comprehensive cognitive assessments, including the Montreal Cognitive Assessment (MoCA) and the Mini-Mental State Examination (MMSE), were administered as part of evaluations at the Emory Cognitive Neurology Clinic, the Emory Goizueta Alzheimer’s Disease Research Center (ADRC), and related studies such as the Emory Healthy Brain Study (EHBS). Diagnostic data were obtained from the ADRC and the Emory Cognitive Neurology Program. Plasma samples were collected using standard National Centralized Repository for Alzheimer’s Disease and Related Dementias (NCRAD) procedures, processed (e.g., 2000 × g for 10 min), and stored in accordance with the 2014 Alzheimer’s Disease Center (ADC) best practices guidelines (39). To quantify plasma pTau217, EDTA plasma samples were prepared according to the manufacturer’s instructions (ALZpath Simoa pTau 217 v2 Assay Kit Quanterix, Billerica, MA, USA). Samples were run in a single batch. Plasma was thawed at room temperature for 45 minutes, centrifuged at 5,000×g for 10 minutes, diluted fourfold, and analyzed on the Simoa HDX platform. Mean intra-assay coefficients of variation (CV) were below 10%. In total, 216 plasma samples from 78 cognitively normal healthy controls, 60 individuals with mild cognitive impairment and 76 AD cases (2 samples analyzed in duplicates) were processed across 3 analytical plates. Plate 3 samples were additionally profiled using the Alamar NULISA CNS panel, including pTau217 measured with the ALZpath antibody. To enable cross-platform integration, both Simoa- and Alamar-derived pTau217 values were transformed into Z-scores and harmonized together for downstream analyses. CSF biomarker positivity was defined for each sample using a cohort-specific tTau/Aβ₁₋₄₂ cutoff (≥ 0.226 = biomarker-positive; < 0.226 = biomarker-negative). Fifteen pooled plasma samples were prepared from corresponding disease groups (5 controls, 5 MCI, and 5 AD). All plasma samples were processed using ENRICHplus kit followed by DIA-MS. Sample information was summarized in **Supplemental Tables 1 and 8**.

### ENRICHplus PreOmics Sample Processing

The ENRICHplus kit (PreOmics, lot# 0000511843) was used for plasma sample processing. The standard manufacturer’s protocol was used with minor changes. First, beads were washed in bulk by adding 1 mL of wash buffer for every 200 µL of bead slurry, shaking at 1,300 rpm for 1 min, and removing the wash buffer using a magnetic stand. This wash step was repeated four times. The washed beads were then pooled and resuspended in binding buffer at twice the original bead slurry volume. Halt™ Protease Inhibitor Cocktail was added to the bead slurry at a final volume equivalent to 0.04× the original bead slurry volume. Next, 50 µL of bead suspension was dispensed into each well of 8-well strips. An equal volume of sample (50 µL) was added and incubated at 30 °C for 30 min with shaking at 1,300 rpm. Following incubation, the supernatant was removed using a magnetic stand. The beads were then washed three times with 100 µL of binding buffer per wash, using the same shaking and magnetic separation steps. For lysis and proteolytic digestion, 40 µL of LYSE buffer was added to each sample, followed by heating at 60 °C for 10 min with shaking. After cooling, 10 µL of enzyme solution was added, and samples were digested overnight at 30 °C with shaking. Following digestion, 60 µL of STOP buffer was added, and samples were loaded onto the kit desalting columns and centrifuged at 1,000 rpm for 1 min. Columns were washed sequentially with 100 µL of WASH 1 buffer and 100 µL of WASH 2 buffer, with each wash centrifuged at 2,250 rpm for 1 min. Peptides were eluted twice with 75 µL of ELUTE buffer and centrifuged at 1,000 rpm for 1 min. The combined eluates were dried by vacuum centrifugation using a SpeedVac.

### Liquid Chromatography and Mass Spectrometry

Each sample was resuspended in 100 µl of loading buffer (0.1% formic acid) and Global Pooled Standard (GPS) samples were prepared by pooling equal amount of peptide from samples within each set. For each sample, 20 µl was loaded onto Evotips and analyzed by liquid chromatography coupled to tandem mass spectrometry. Each sample contained an equivalent amount of Biognosys iRT standards, a set of 11 synthetic, non-naturally occurring peptides, spiked into every sample to enable consistent retention time normalization and LC–MS quality control. Peptide eluents were separated on IonOpticks column (15 cm × 75 µm internal diameter (ID) packed with 1.7µm resin) by Evosep One (Evosep). Buffer A was water with 0.1% (vol/vol) formic acid, and buffer B was 100% (vol/vol) acetonitrile in water with 0.1% (vol/vol) formic acid. Elution was performed using the preset 40 samples per day (SPD) Whisper Zoom method. Peptides were analyzed on an Orbitrap Astral mass spectrometer (Thermo Fisher Scientific) equipped with a FAIMS Pro high-field asymmetric waveform ion mobility spectrometry source (Thermo Fisher Scientific). A single FAIMS compensation voltage (CV) of −40 V was used. Each acquisition cycle consisted of one full MS1 scan acquired over an m/z range of 380–980 at a resolution of 240,000, with an automatic gain control (AGC) target of 500% and a maximum injection time of 3 ms. Higher-energy collisional dissociation (HCD) data-independent acquisition (DIA) scans were collected across the full precursor range of 380–980 m/z using 3 m/z isolation windows, a cycle time of 0.6 s, and a maximum injection time of 3 ms. The normalized collision energy was set to 27%, and fragment ions were acquired over an m/z range of 150–2,000.

### Database Search

Spectronaut (version 19.9.250512.62635) was used to search all raw files in default library free mode using a human UniProt database (downloaded February 2019 and supplemented with amyloid-beta species and APOE variant isoforms). All parameters were kept at default.

### Data Normalization and Variance Correction

The Spectronaut outputs from the three plates searched together (n = 216), Plate 3 searched independently (n = 36), and the pooled plasma samples (n = 15) were each generated separately. Raw protein intensity values were first normalized by total MS signal, scaling each sample’s summed abundance to the maximum total signal observed across all samples. This procedure minimizes sample-to-sample variation due to differences in total protein load or instrument response. Proteins with more than 50% missing values were then removed from each dataset prior to downstream analysis (no imputation of missing values was performed). A tunable median polish approach (TAMPOR) mode 4 was used to adjust technical variance within each dataset as previously described

(40). The algorithm is fully documented and available as an R function, which can be downloaded from https://github.com/edammer/TAMPOR. Finally, the iRT peptide measurements and the duplicate samples were removed from the matrix before downstream analysis. To ensure consistent protein-level representation during downstream analysis, protein groups annotated with multiple gene symbols and UniProt accessions were collapsed to a single representative entry by retaining the first gene symbol and corresponding accession.

### Proteome Overlap and Correlations across Individual Proteomic Platforms

Proteome overlap across individual platforms was visualized using the *venneuler* R package (v1.1.4). To evaluate cross-platform consistency, we compared the microbead-enriched DIA-MS data (PreOmics DIA-MS) with plasma measurements obtained using heparin enrichment followed by TMT-MS (Heparin TMT-MS) and SomaScan® aptamer-based technology (SomaLogic, Colorado, USA). Heparin TMT-MS was performed on the same 36 samples in Plate 3, as described above. SomaScan data were available for 35 of these individuals (control = 18, AD = 17) and were obtained from prior publications (29, 41). Correlation analyses of the log2 fold-change (AD vs. Control) were carried out and visualized employing the *verboseScatterplot* function from the R WGCNA package, utilizing the Pearson correlation coefficient and Student’s *p*-values to determine the statistical significance.

### Weighted Gene Co-Expression Network Analysis (WGCNA)

Network analysis was performed using the WGCNA R package (v1.72.5), as previously described (29, 33–35). A weighted protein co-expression network was constructed from the PreOmics DIA-MS dataset using the variance corrected and log₂-transformed abundance matrix (5,823 proteins × 214 samples). After removal of 6 connectivity outliers and applying a 50% missing-value threshold, the final input matrix included 5,811 proteins across 208 samples. The network was generated using the *blockwiseModules()* function with the following parameters: soft-threshold power = 10, deepSplit = 4, minimum module size = 10, merge cut height = 0.07, signed network type, partitioning around medoids (PAM) respecting the dendrogram, mean TOM denominator, and a reassignment threshold of *p* < 0.05. Clustering was performed within a single block, resulting in 29 distinct protein co-expression modules (≥ 10 proteins each). To ensure robust module assignments, a post hoc cleanup procedure was applied to refine the kME (module membership) table. Proteins with intramodular kME < 0.30 were first removed. Subsequently, unassigned (“gray”) proteins with a maximum kME > 0.30 were reassigned to the corresponding module. Proteins with an intramodular kME more than 0.10 lower than their maximum kME to another module were also reassigned. Module eigenproteins (MEs) and signed kME values were recalculated using *moduleEigengenes()* and *signedKME()* functions, respectively. This iterative refinement continued until no further reassignments improved consistency (five iterations in total for the plasma network), reducing unassigned proteins from 2,267 (39.0%) to 1,301 (22.4%). Module correlations to clinical and demographic traits, as well as biomarker measurements were evaluated with biweight midcorrelation (bicor) analysis using *WGCNA::bicorAndPvalue()* function. Eigenprotein differential abundance was analyzed with one-way ANOVA across groups.

### Gene Ontology (GO) Analysis

All functional enrichment analyses were performed using the *GOparallel* package (available at https://github.com/edammer/GOparallel). Gene Ontology annotations were retrieved from the Bader Lab’s monthly updated .GMT files (36). Enrichment significance was assessed using a one-tailed Fisher’s exact test (FET) followed by Benjamini-Hochberg (BH) false discovery rate (FDR) correction, and both Z-scores and *p*-values were calculated. A cutoff of Z-score > 1.96 (BH FDR corrected *p* < 0.05 and a minimum of five genes per ontology) was used as filter prior to pruning the ontologies.

### Protein Differential Abundance and Bicor Volcano Plot

For pairwise differential abundance, protein abundance was compared between groups using Student’s *t*-test. Volcano plots (x = log₂ fold-change; y = −log₁₀ *p* values) were generated in R (v4.3.1) with the ggplot2 package. For biomarker–protein association (bicor volcano), we computed biweight midcorrelation (bicor) between each protein’s abundance and AD biomarkers (CSF Aβ₁₋₄₂, CSF tTau, CSF pTau181, CSF tTau/Aβ₁₋₄₂ ratio, and plasma pTau217). Bicor and *p*-values were calculated in R using *WGCNA::bicorAndPvalue()* function. Bicor volcano plots displayed bicor (r) in x-axis and −log₁₀ *p* values in y-axis. Each significance was defined at *p* < 0.05.

### Protein-Biomarker Association Analysis and Visualization

Protein-biomarker associations were assessed using linear regression models relating individual plasma protein abundances to established Alzheimer’s disease-related CSF and plasma biomarkers. Biomarkers examined included CSF Aβ₁₋₄₂, CSF tTau, CSF pTau181, CSF tTau/Aβ₁₋₄₂ ratio and plasma pTau217, all expressed as Z-scores. Analyses were restricted to participants with no missing values for all biomarkers of interest (*n* = 129). For each protein-biomarker pair, an independent linear regression model was fitted with protein abundance as the dependent variable and the biomarker Z-score as the independent variable. Models were fitted using ordinary least squares. The regression coefficient corresponding to the biomarker term and its associated two-sided *p*-values were extracted from each model. To facilitate comparison of effect sizes across proteins and biomarkers, standardized regression coefficients were calculated post hoc by rescaling the estimated regression coefficient using the ratio of the standard deviation of the biomarker to the standard deviation of the protein abundance. These standardized coefficients reflect the expected change (in standard deviation units) of protein abundance per one standard deviation change in the biomarker. All analyses were performed in R.

For visualization purposes, proteins were ranked separately for each biomarker based on nominal *p*-values derived from the linear regression models. The top 25 proteins per biomarker were selected, and the top 80 of the union of these proteins across all five biomarkers was displayed in the final visualization. Regression coefficients and *p*-values for these proteins were retained for all biomarkers, enabling direct comparison across biomarker modalities. Associations between proteins and biomarkers were visualized using a dot-plot representation. Regression coefficients were displayed using a diverging color scale centered at zero, with color intensity reflecting the direction and magnitude of association. Statistical significance was encoded by point size, proportional to the −log₁₀ *p*-values. Proteins were ordered along the x-axis according to a predefined sequence reflecting biological groupings derived from network analyses, and axis label colors were customized to indicate module membership. Biomarkers were displayed on the y-axis as discrete categories. Visualization was implemented using ggplot2.

### Top Ten Protein Selection and Group Classification Analysis

To classify subject groups, we selected the top ten plasma proteins from a set of 1,763 targets quantified without missing values across 214 samples. We restricted the analysis to proteins with complete data across all samples because the modeling framework did not permit missing values, and we sought to avoid potential biases introduced by data imputation or uneven protein completeness. Feature selection was performed using Recursive Feature Elimination (RFE) with a linear Support Vector Classifier (SVC) to identify the ten most informative proteins that best distinguish CSF biomarker–positive from biomarker–negative groups. SHAP (SHapley Additive exPlanations) analysis was then conducted in Python (v3.12.10) using the same svm.SVC classifier and the Kernel Explainer to evaluate the contribution of each selected protein to the classification model. Hyperparameters for the SVM were optimized using the *optuna.create_study()* function. Receiver operating characteristic (ROC) analysis was subsequently performed in R (v4.3.1) to evaluate classification performance. A generalized linear model (binomial family) was fitted to the selected protein measurements (plasma pTau217 alone, the selected 10-protein panel, and the combination of both) for binary group classification. ROC curves, areas under the curve (AUCs), and 95% confidence intervals were computed using the *pROC* package (42).

### Over Representation Analysis

The overlap between plasma and brain co-expression modules established previously (29) was assessed by overrepresentation analysis (ORA) using one-tailed Fisher’s exact tests (FET). Enrichment significance was assessed by extracting the *p*-values from FET followed by Benjamini–Hochberg (BH) false discovery rate (FDR) correction. Those modules with BH FDR-corrected *p* < 0.05 were considered significant. The background for this analysis is the total number of proteins quantified in both datasets.

### Cell-Type and Human Organ Marker Enrichment Analysis

Cell-type enrichment for each brain module was performed as previously described (29, 43). Briefly, the corresponding gene symbols of each module were cross-referenced with cell-type–specific gene lists derived from previously published RNA-seq data (15, 44). Significance of cell-type enrichment within each module was then determined using a one-tailed Fisher’s exact test followed by BH FDR correction. The algorithm is fully documented and available as an R function, which can be downloaded from https://github.com/edammer/CellTypeFET. Human organ enrichment for plasma network modules was evaluated by testing the overlap between module proteins and organ-specific marker gene sets (13). Significance of organ enrichment was determined using FET followed by BH FDR correction.

### Module Preservation and Cross-Network Comparison

Preservation of plasma co-expression modules within the consensus human brain proteomic network (29) was evaluated using the *WGCNA::modulePreservation()* function, as previously described (15, 43, 45), using the plasma network as template. Modules with less than 35 proteins (M27, M28, M29) were removed and considered to be not preserved. To assess module-wise concordance in AD-related changes across plasma and brain, we compared the log₂ fold-change of module eigenproteins in plasma network (AD/MCI vs. CTL) versus the synthetic eigenproteins derived from the brain network (AD vs. CTL). Only individuals classified as AD or MCI with CSF tTau/Aβ₁₋₄₂ ratio ≥ 0.226 and controls with CSF tTau/Aβ₁₋₄₂ ratio < 0.226 were included. Synthetic brain modules were constructed by selecting proteins (minimum 4 proteins for each module) overlapping with each plasma module and representing the top 20% of hub proteins ranked by intramodular connectivity (kME). The first principal component of these proteins was computed using the *moduleEigengenes()* function in WGCNA, yielding synthetic eigenprotein values as described previously (15, 43, 45). Protein-level comparison was performed by comparing the log₂ fold-change of protein abundances in plasma (AD/MCI vs. CTL) and brain (AD vs. CTL) datasets.

### Other Analysis

Heatmap of correlations between CSF and plasma biomarkers were computed and visualized using *ggcorrplot()* function (method = “square”) in R (v4.3.1). Pairwise correlations were further assessed using the *cor.test()* function and displayed as scatter plots generated with the *plot()* function in R (v4.3.1). Potential plasma sample contamination by platelets, erythrocytes, and coagulation-related proteins was assessed using the Baize software (46).

### Disclosures

N.T.S, A.I.L and D.M.D. are co-founders and consultants of Emtherapro. D.M.D. and N.T.S are co-founders of Arc Proteomics. N.T.S is a co-founder of StitchRx.

### Data Availability

Raw mass spectrometry files, Spectronaut search and quantification results, pre- and post-normalization protein abundance data, and associated metadata related to this manuscript are available at: https://www.synapse.org/Synapse:syn74967843.

## Results

### Deep Plasma Proteomics Reveals Broad and Reproducible Coverage in a Well-Characterized AD Cohort

To test whether microbead-enriched plasma proteomics captures biologically meaningful AD signals and reflects brain pathology, we applied an ENRICHplus microbead-based workflow with Orbitrap Astral DIA-MS to a well-characterized cohort with both plasma and CSF AD biomarker measurements. Participants were phenotyped with annual assessments at the Emory Goizueta Alzheimer’s Disease Research Center (ADRC), with clinical diagnoses assigned by consensus conference. A total of 214 individuals were analyzed, including 78 cognitively normal controls, 60 with mild cognitive impairment (MCI), and 76 with AD (**Figure 1A**). Clinical and biomarker trait distributions, including MMSE, CSF Aβ₁₋₄₂, total tau (tTau), pTau181, tTau/Aβ₁₋₄₂ ratio, and plasma pTau217, showed expected disease-related differences (**Figure 1B**). Of the 214 participants, 129 had complete CSF AD biomarker and plasma pTau217 data (**Figure 1C**). Plasma pTau217 correlated significantly with the CSF tTau/Aβ₁₋₄₂ ratio, tTau and pTau181 (**Figure 1D; Supplemental Figure 1**).

**Figure 1.**
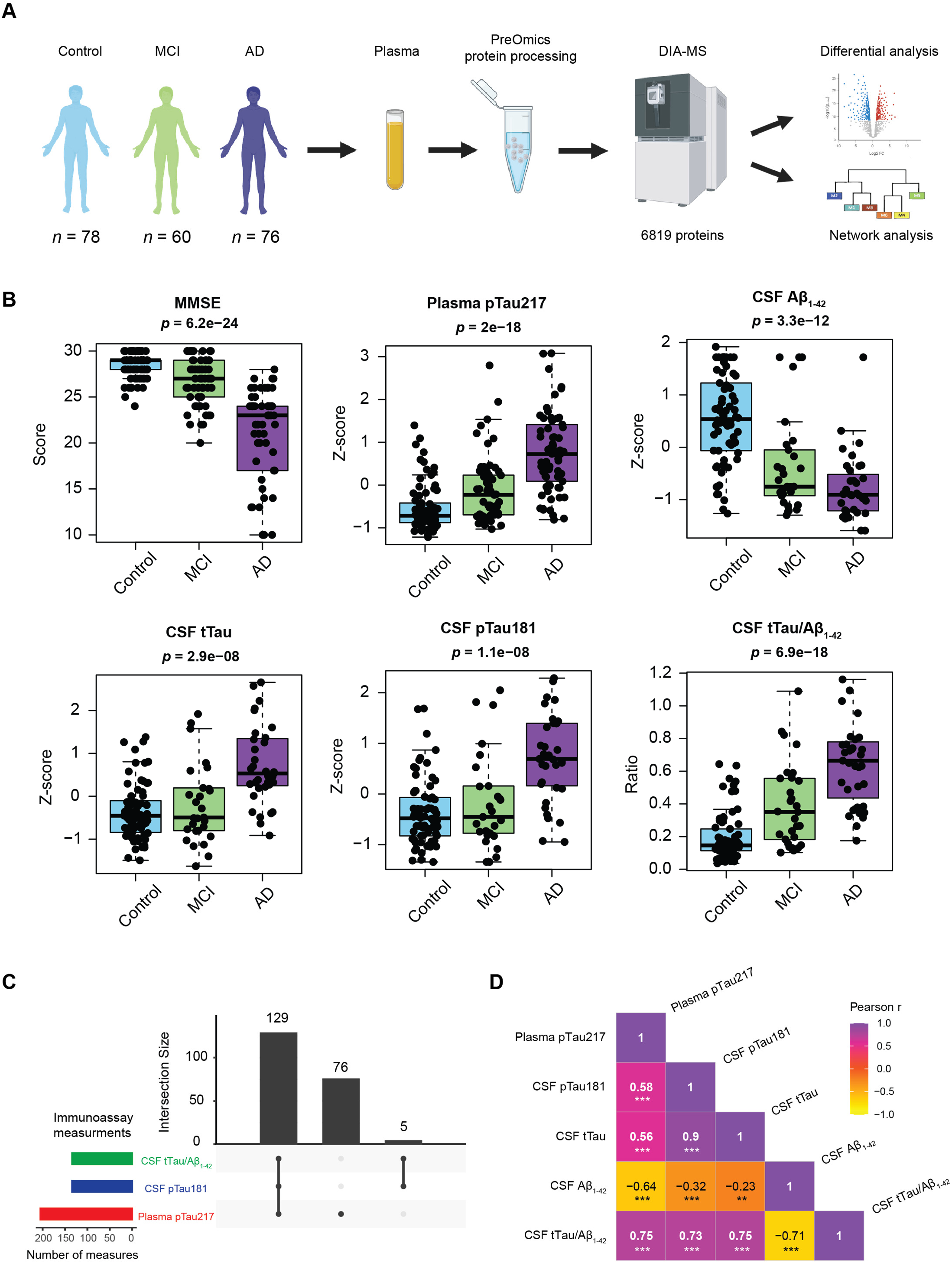
Overview of the Plasma DIA-MS Workflow, Clinical Cohort, and AD Biomarker Characterization. **A)** A total of 216 plasma samples obtained from 78 Control, 60 MCI, and 76 AD (2 duplicate samples) underwent ENRICHplus microbead-based processing (from PreOmics) followed by data-independent acquisition mass spectrometry (DIA-MS) on an Orbitrap Astral mass spectrometer, identifying 6,819 proteins in total. **B)** Distribution of AD-related traits, including cognition (MMSE score), CSF Aβ1-42, CSF tTau, CSF pTau181, CSF tTau/Aβ1-42 ratio, and plasma pTau217, shown for all unique cases (78 Control, 60 MCI, and 76 AD). The statistical significance of group differences was assessed using one-way ANOVA, where *p* < 0.05 was considered significant. **C)** Number of samples with immunoassay measurements: 129 samples had complete measurements for CSF Aβ1-42, CSF tTau, CSF pTau181, CSF tTau/Aβ1-42 ratio, and plasma pTau217; 76 samples had plasma pTau217 only, and 5 samples had CSF measures only. **D)** Pearson correlation matrix between Z-scored plasma pTau217 and CSF biomarkers (tTau/Aâ1-42 and Z-scored Aβ1-42, tTau, and pTau181). Purple indicates positive correlations and yellow indicates negative correlations. Significance levels are denoted by overlaid asterisks (\*\**p* < 0.01, \*\*\**p* < 0.001). Strong positive correlations were observed between plasma pTau217 and CSF tTau/Aβ1-42 ratio (*r* = 0.75).

Following ENRICHplus microbead enrichment of 216 samples (including 2 runs in duplicate), DIA-MS was performed and instrument performance was monitored using 12 Global Pooled Standard (GPS) samples, each containing equal peptide amounts from all samples, interspersed across three analytical plates (**Supplemental Figure 2A**). Overall, we identified 6,819 proteins (65,045 unique peptides) across all samples and GPS (**Supplemental Table 2**); 1,763 were quantified without missing values and 5,823 were detected in >114 samples (**Supplemental Figure 2B**). These reliably measured 5,823 proteins spanned ∼10 orders of magnitude in plasma abundance, reaching concentrations as low as 2.90 pg/mL, including low-abundance proteins such as SYNJ1 and PLCB4 (**Supplemental Figure 2C, Supplemental Table 3**) (47). Prior to data normalization, we assessed plasma sample quality using Baize software (46) to estimate potential contamination from platelets, erythrocytes, and coagulation-related proteins. According to the interpretation guidelines provided in the original report (46), erythrocyte contamination was negligible (mean index = 0.0248) and coagulation-related contamination was minimal (mean index = 14.3), while platelet contamination was in the low-to-moderate range (mean index = 0.0196) (**Supplemental Figure 2D, Supplemental Table 4).** This level of platelet-associated carryover is consistent with the plasma collection protocol used in this study, which employs a single centrifugation step (2,000 × g for 10 min) rather than more stringent procedures that further reduce platelet carryover (46). These results indicate generally good plasma sample quality and suggest that pre-analytical contamination was unlikely to materially influence the downstream proteomic analyses.

To normalize and adjust for covariates, we applied sample-wise normalization followed by median normalization to the protein abundance matrix. Before median normalization (**Supplemental Table 5**), multidimensional scaling (MDS) plots showed modest plate-driven clustering, whereas after correction, samples clustered uniformly, indicating effective removal of plate effects (**Supplemental Figure 3A, Supplemental Table 6**). Variance partitioning confirmed that plate effects were substantially reduced after normalization, with age, sex, race, and diagnosis explaining a greater proportion of the remaining variance (**Supplemental Figure 3B**). Technical reproducibility assessed by GPS was high, with > 5,300 proteins quantified without missing values and a median post-normalization CV < 15% across GPS samples (**Supplemental Figure 4, Supplemental Table 7**). Consistent with this, the microbead enrichment workflow showed strong reproducibility, as 15 pooled plasma samples processed the same way as the cohort samples yielded > 6,200 proteins with a median post-normalization CV of ∼17% (**Supplemental Figure 5, Supplemental Tables 8–12**). This represents only a ∼2% increase relative to technical CVs, indicating that sample processing introduces minimal additional variability beyond MS measurement. These data demonstrate broad, consistent plasma proteome coverage across control, MCI, and AD, supporting downstream analyses of disease-associated molecular variation.

### Cross-Platform Comparison of AD-Associated Plasma Proteome Signatures

To evaluate the consistency of AD-associated protein changes identified by microbead-enriched DIA-MS, we compared results from a subset of overlapping control and AD plasma samples profiled across two independent proteomic platforms: SomaScan® (n = 35) and heparin-enriched TMT-MS (n = 36) (**Supplemental Figure 6**). All datasets were median-normalized, and analyses were restricted to proteins quantified in ≥18 samples per platform (**Supplemental Tables 13-15**). Across all three platforms, 9,373 unique plasma proteins were represented, with SomaScan profiling the greatest number (*N* = 6,374), followed by microbead-enriched DIA-MS (PreOmics DIA-MS, *N* = 5,295) and heparin-enriched TMT-MS (Heparin TMT-MS, *N* = 2,077) (**Supplemental Figure 6A, Supplemental Table 16**). Notably, PreOmics DIA-MS quantified 2,803 proteins not detected by SomaScan, including proteins enriched for mitochondrial and structural pathways, highlighting complementary coverage relative to affinity-based assays (**Supplemental Figure 6B**).

We next assessed cross-platform agreement in AD-associated plasma changes by correlating log₂ fold-changes (AD vs. control) for overlapping proteins significant in both datasets (*p* < 0.05) (**Supplemental Figure 6C–D, Supplemental Tables 17-19)**. Effect sizes were highly concordant between PreOmics DIA-MS and Heparin TMT-MS (*n* = 391; *r* = 0.78, *p* = 3.3 × 10⁻⁸¹), as well as between PreOmics DIA-MS and SomaScan (*n* = 531; *r* = 0.70, *p* = 2.2 × 10⁻⁷⁹). Directionality of change was also highly consistent across platforms, with 91% and 82% concordance for microbead-enriched DIA-MS relative to heparin-enriched TMT-MS and SomaScan, respectively. The remaining discrepancies potentially reflect TMT-MS ratio compression in the heparin dataset and aptamer-specific binding biases in the SomaScan dataset. Proteins showing consistent increase in AD across platforms included ECM/matrisome markers (SMOC1, GPNMB, HTRA1, CTHRC1, SPON1, FRZB) and metabolic/inflammatory proteins (GIP, BGN, CLU, PLA2G7, ANGPTL8, GCG, LPL), while UBA6 and BZW2 showed consistent decreases (**Supplemental Figure 6E**). Together, these results support strong cross-platform reproducibility of AD-associated plasma protein changes and highlight the complementary proteome coverage provided by microbead enrichment relative to affinity-based methods.

### Plasma proteomic remodeling across AD progression reveals pathways linked to cognition and aging

To determine whether plasma proteomic alterations reflect disease progression and cognitive decline, we performed differential abundance and correlation analyses across diagnostic groups (control, MCI, and AD) (**Figure 2**). These analyses aimed to identify plasma proteins that change with clinical stage and to assess whether these changes are linked to cognitive impairment or aging. Differential abundance analysis revealed widespread plasma proteomic differences across disease stages (**Figure 2A, Supplemental Table 20**). Compared to controls, 1,257 proteins were altered in MCI and 2,018 proteins in AD, while 331 proteins differed between AD and MCI. The increased number of differentially abundant proteins in AD relative to MCI is consistent with greater disease severity and parallels patterns previously observed in proteomic analyses in brain tissue across clinical and pathological stages of AD (43, 45, 48). Proteins implicated in lipid metabolism, inflammation, and vascular biology were among the most significantly altered, including APOE, APOD, PLA2G7, SMOC1, and FRZB (29, 45, 49), as well as erythrocyte-associated proteins such as GYPA, RHCE, and SPTA1 (50–52), suggesting systemic metabolic and hematologic changes accompanying disease progression. To assess associations with cognitive performance, we next examined relationships between plasma proteins and MMSE. A total of 1,067 proteins were significantly correlated with cognition, including 620 positively and 447 negatively associated proteins (**Figure 2B, Supplemental Table 21**). Proteins negatively associated with MMSE were significantly enriched for extracellular matrix and structural pathways, including keratinization and cytoskeletal organization, whereas proteins positively associated with MMSE were enriched for mitochondrial metabolism, oxidative phosphorylation, and tricarboxylic acid cycle pathways, suggesting reduced mitochondrial-associated protein abundance with worsening cognition (**Figure 2D**). Correlation analysis with age revealed distinct plasma proteomic signatures associated with aging (**Figure 2C and 2E, Supplemental Table 22**). Proteins that increased with age included RELN, EFEMP1, and SVEP1, which are involved in extracellular matrix and tissue remodeling pathways, whereas proteins such as IMMT, SQOR, and ITPR2, associated with mitochondrial metabolism and intracellular signaling, were negatively correlated with age. These results demonstrate that plasma proteomic alterations across AD progression capture biological signatures of mitochondrial dysfunction, extracellular remodeling, and systemic aging, linking circulating proteins to cognitive decline and disease severity.

**Figure 2.**
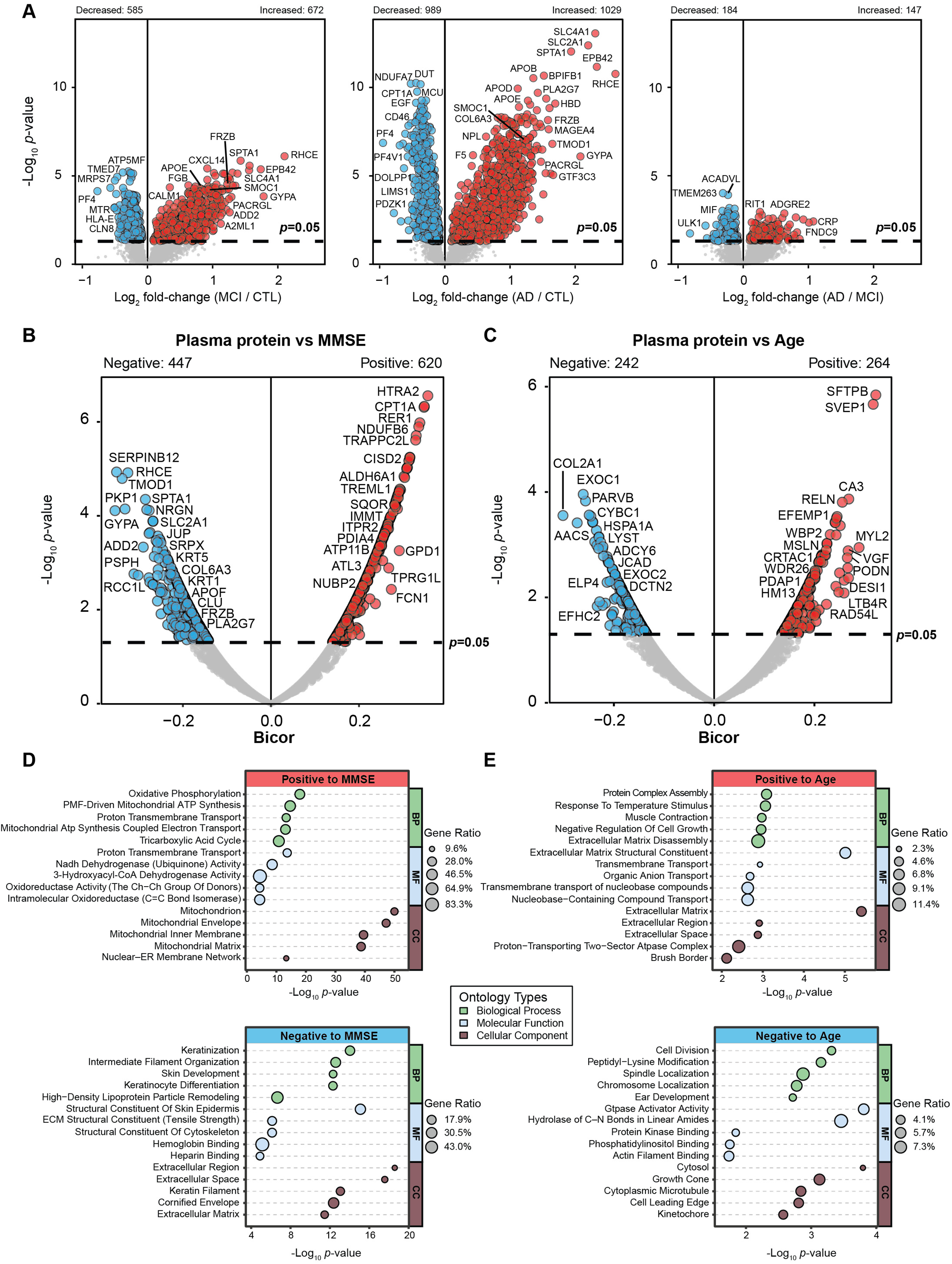
Plasma Proteomic Changes Across Clinical Stage, Cognition, and Aging. **A)** Volcano plots showing differential protein abundance across diagnostic groups (MCI vs. CTL, AD vs. CTL, and AD vs. MCI). The x-axis represents log₂ fold-change and the y-axis shows –log₁₀ *p*-values. Proteins passing the significance threshold (*p* < 0.05, dashed horizontal line) are colored as red (increased) or blue (decreased), with representative proteins labeled. **B-C**) Volcano plots showing biweight midcorrelations (bicor) between plasma protein abundance and MMSE (left) or age (right). The x-axis shows bicor and the y-axis represents –log₁₀ *p*-values. Proteins passing the significance threshold (*p* < 0.05, dashed horizontal line) are highlighted as red (positive correlations) or blue (negative correlations). **D-E**) Gene Ontology (GO) enrichment of proteins positively or negatively associated with MMSE (left) and age (right), with *p-*values < 0.05. Dot size reflects gene ratio and color indicates enrichment significance (–log₁₀ *p*-value). Abbreviations: Mild cognitive impairment (MCI), cognitively normal controls (CTL) and Alzheimer’s disease (AD).

### Plasma Proteome Network Reveals Modules Linked to AD Pathology and Demographic Traits

Given the depth of the microbead-enriched proteome coverage across the 214 plasma samples, we first assessed whether the plasma proteome could be organized into biologically meaningful clusters and whether these clusters related to clinical and pathological features of AD. Using Weighted Gene Co-expression Network Analysis (WGCNA) on 5,823 reliably measured proteins, we identified 29 co-expression modules (**Figure 3A, Supplemental Table 23**). Gene Ontology (GO) enrichment analysis showed that these modules represent diverse biological processes, including lipid metabolism, extracellular matrix organization, immune response, mitochondrial function, and proteostasis. We next examined associations between module eigenproteins and demographic, clinical, and biomarker traits. As shown in **Figure 3B–C**, several modules displayed strong biweight midcorrelations (bicor) with age, sex, race, APOE4 gene dose, MMSE, and CSF or plasma AD biomarkers (CSF Aβ₁₋₄₂, tTau, pTau181, tTau/Aβ₁₋₄₂, and plasma pTau217). For example, M1 (LDL-cholesterol/matrisome), M2 (ECM/immune response), and M26 (keratin) exhibited the strongest positive correlations with AD-related traits and APOE4 gene dose, along with notable associations with sex and race. In contrast, multiple mitochondrial and proteostasis-related modules (M3, M4, M5, M6, M8, M11, M12) showed reduced abundance in AD, consistent with impaired metabolic and bioenergetic processes in both brain and CSF (45, 53).

**Figure 3.**
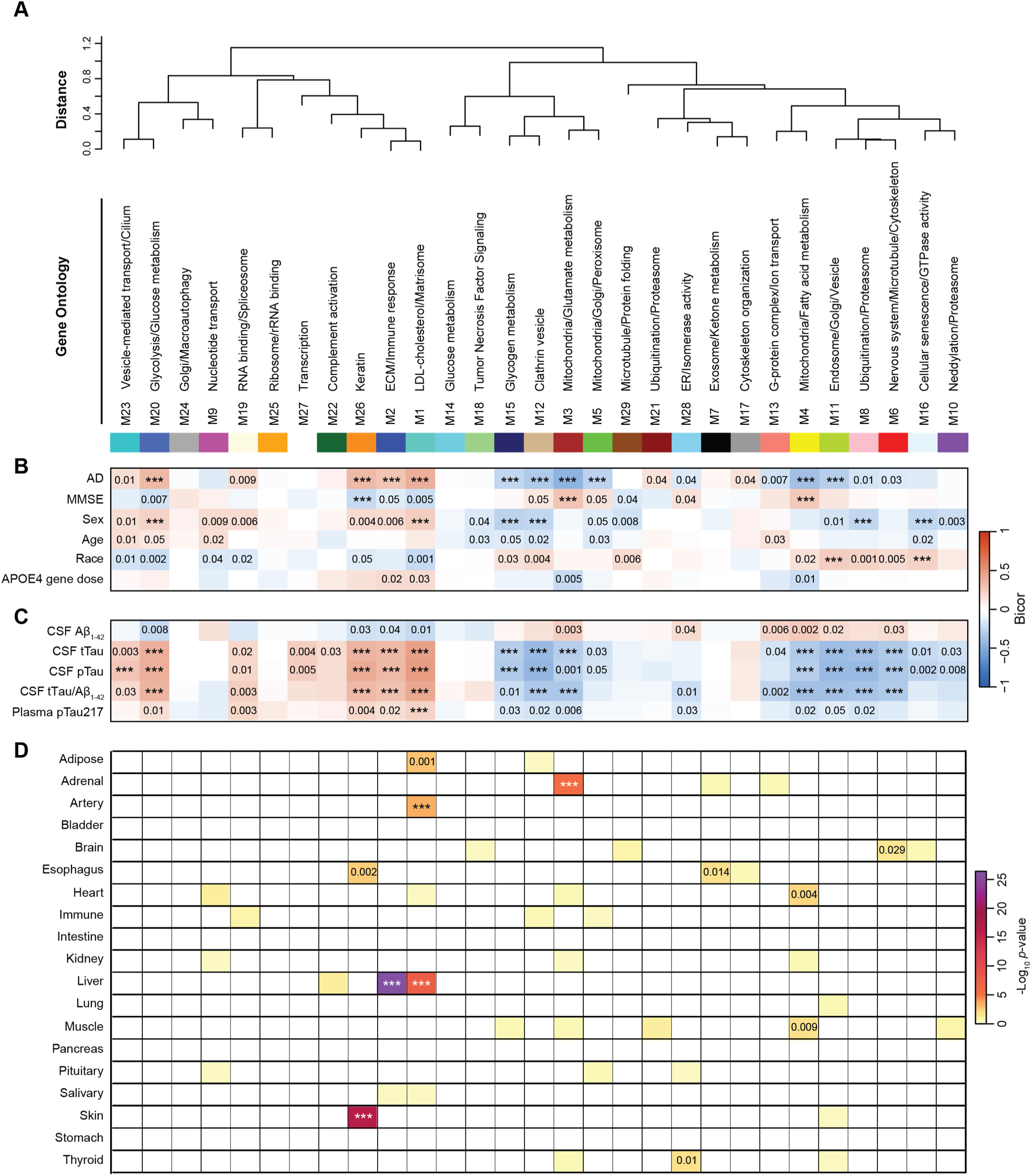
Systems-Level Plasma Proteome Network Analysis Reveals AD-Associated Biological Pathways and Tissue Enrichment. **A)** A protein co-expression network identified 29 distinct protein modules (M1-M29). Module relationships are shown in the dendrogram, and Gene Ontology (GO) enrichment analysis highlights the predominant biological processes represented in each module. **B–C**) The heatmap displays biweight midcorrelations (bicor) between module eigenproteins and clinical or biological traits, including AD status, age, sex, race, APOE4 gene dose, MMSE cognitive score, and biofluid biomarkers (CSF Aβ1-42, CSF tTau, CSF pTau181, CSF tTau/Aβ1-42 ratio, and plasma pTau217). Red indicates positive correlations, and blue indicates negative correlations. Statistical significance (Student’s *t*-test) is indicated by overlaid numbers or asterisks (\*\*\**p* < 0.001). Modules M1, M2, and M26 showed the strongest positive correlations with AD pathology, whereas mitochondrial and proteostasis-related modules (M3, M4, M5, M6, M8, M11and M12) exhibited decreased abundance in AD plasma. **D**) Module tissue enrichment was determined using Fisher’s Exact Test (FET). The organ origin of each module was inferred by testing the overlap between module proteins and human tissue-specific markers (adipose, adrenal, artery, bladder, brain, esophagus, heart, immune, intestine, kidney, liver, lung, pancreas, pituitary, salivary gland, skin, stomach, and thyroid). The intensity of color shading reflects the strength of enrichment, and significance is denoted by overlaid numbers or asterisks (\*\*\**p* < 0.001). All FET-derived *p*-values were corrected using the Benjamini–Hochberg false discovery rate (FDR).

### Plasma Protein Modules Map to Distinct Tissue Origins

To define the potential biological origins of plasma protein modules, we tested whether module members were enriched for tissue-specific gene signatures across 17 human organs (13). Because circulating proteins derive from multiple sources, including liver, vasculature, immune cells, and brain, this analysis provides important context for interpreting disease-associated changes and helps distinguish systemic signals from those reflecting brain or vascular processes relevant to AD. Several plasma modules showed strong enrichment for proteins preferentially expressed in specific tissues (**Figure 3D**). Modules associated with extracellular matrix and immune pathways, including M1 (LDL-cholesterol/matrisome) and M2 (ECM/immune response), were significantly enriched for arterial and vascular tissue signatures, consistent with the prominent contribution of vascular and extracellular proteins to the circulating proteome. Modules involved in metabolic pathways, such as M20 (glycolysis/glucose metabolism), showed enrichment for liver-associated proteins, reflecting the central role of hepatic secretion in shaping the plasma proteome. In contrast, M26 (keratin) showed strong enrichment for skin-derived proteins, consistent with the presence of keratin and epithelial structural proteins commonly detected in plasma. Importantly, M6 (nervous system/microtubule/cytoskeleton) showed enrichment for brain-related signatures and was among the modules most strongly associated with CSF AD biomarkers, suggesting that a subset of circulating proteins may reflect CNS biology. These findings indicate that plasma proteomic networks integrate proteins originating from multiple tissues, providing a framework for interpreting how circulating protein signatures may capture both systemic and neurodegenerative processes in AD.

### AD-Associated Plasma Protein Modules Vary by Sex and Race

Prior CSF proteomic network analyses have shown that module structure and abundance differ by sex and race (54, 55), with these factors interacting with disease status and core AD biomarkers. We therefore evaluated how plasma module abundance varied across diagnosis, sex, and race. Eight modules showing significant differences across control, MCI, and AD groups were presented in **Figure 4**. As an example, modules significantly positively associated with AD, including M1 (LDL-cholesterol/matrisome), M2 (ECM/immune response), M20 (glycolysis/glucose metabolism), and M26 (keratin), showed stepwise increases from control to MCI to AD. These increases were more pronounced in males and in non-Hispanic White (NHW) individuals (**Figure 4A**). Conversely, modules decreased in AD, including M12 (clathrin vesicle), M4 (mitochondria/fatty acid metabolism), M8 (ubiquitination/proteasome), and M11 (endosome/Golgi/vesicle), tended to be higher in females and in African American (AA) individuals (**Figure 4B**). Collectively, these results show that the plasma proteome is organized into coherent, co-regulated modules linked to biological pathways and AD-related clinical differences. Module abundance also varies by sex and race, indicating that plasma network signatures reflect an interplay between disease biology and demographic factors.

**Figure 4.**
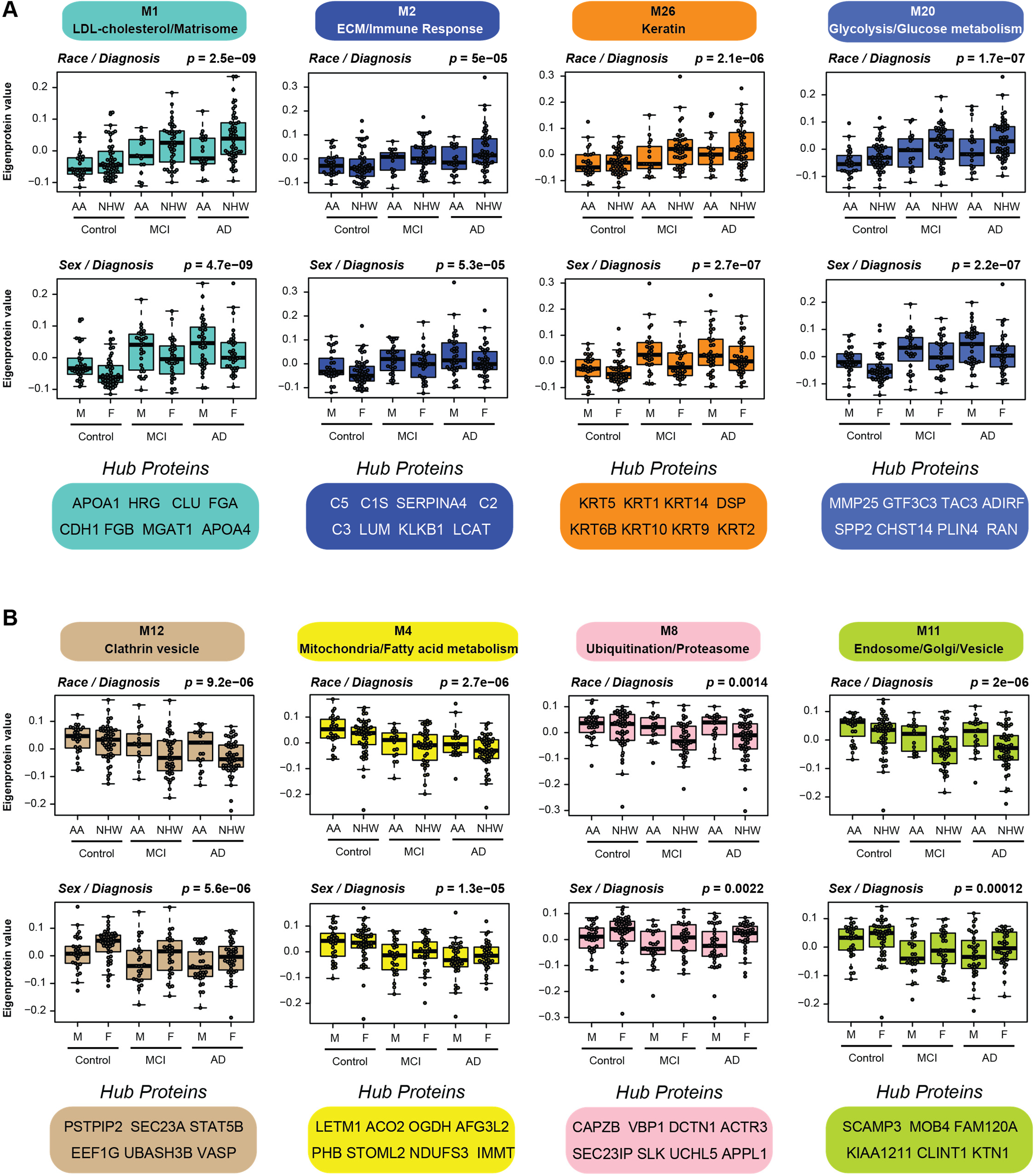
Plasma Network Module Differences Across Sex, Race, and AD Diagnosis. **A-B)** Protein abundance trends across diagnosis stratified by race and sex are shown for 8 of the 29 plasma network modules significantly correlated with AD-related traits. Module abundance is represented by eigenprotein values derived from the plasma network (Control = 78, MCI = 60, AD = 76). Statistical significance of group differences was assessed using one-way ANOVA (*p* < 0.05). Modules **M1** (LDL-cholesterol/matrisome), **M2** (ECM/immune response), **M26** (keratin), and **M20** (glycolysis/glucose metabolism) displayed increased protein abundance in AD plasma, with higher levels in Non-Hispanic White compared to African American individuals and in males compared to females. In contrast, **M12** (clathrin vesicle), **M4** (mitochondria/fatty acid metabolism), **M8** (ubiquitination/proteasome), and **M11** (endosome/Golgi/vesicle) showed decreased abundance in AD plasma, with lower levels in Non-Hispanic White compared to African American individuals and in males compared to females.

### Association of Plasma Proteome Signatures with CSF AD Biomarkers

CSF Aβ₁₋₄₂, pTau181, and tTau/Aβ₁₋₄₂ ratio are well established AD biomarkers used to define amyloid and tau pathology (AT status), whereas plasma pTau species, particularly pTau217, serve as minimally invasive markers of amyloid burden (56, 57). To determine whether plasma proteomic changes associate with AD biomarker status, we analyzed the 129 individuals with both CSF tTau/Aβ₁₋₄₂ ratio and plasma pTau217 measurements and integrated these immunoassay markers with our plasma proteomic network. CSF biomarker positivity was defined using a cohort-specific tTau/Aβ₁₋₄₂ cutoff. As expected, CSF tTau/Aβ₁₋₄₂ ratios differed markedly across diagnostic groups (*p* = 1.1 × 10⁻¹⁷) (**Figure 5A**). Differential abundance analysis revealed widespread proteomic changes, with 810 of the 5,811 network proteins increased and 763 decreased in biomarker-positive individuals (**Figure 5B, Supplemental Table 24**). Proteins elevated in CSF biomarker-positive individuals clustered in modules positively associated with AD-related traits (M1, M2, M26), whereas proteins reduced in biomarker-positive subjects were concentrated in mitochondrial and proteostasis-related modules that are decreased in AD (M3, M4, M5, M6, M8, M11, M12). Notably, those increased proteins in M1 and M2 included matrisome-associated proteins involved in amyloid pathology and cerebral amyloid angiopathy (CAA), such as FRZB, APOE, SMOC1, CLU, SPON1, CD14, BGN, PROS1, DPP4, and NAMPT (15, 58).

**Figure 5.**
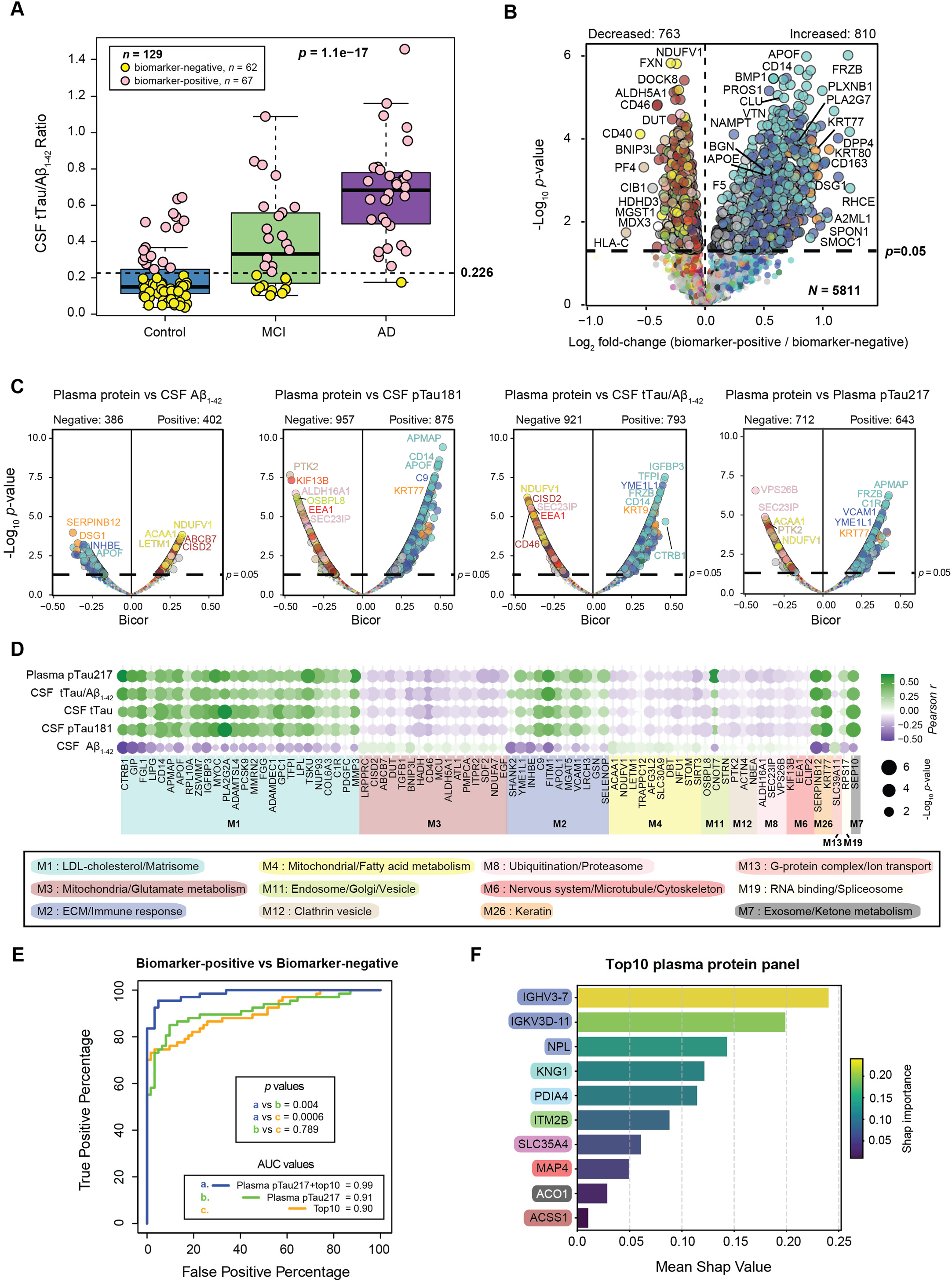
Plasma Proteomic Signatures Associated with CSF Biomarkers in AD. **A)** CSF tTau/Aâ₁₋₄₂ ratios across 129 samples with both plasma pTau217 and CSF biomarker measurements. Group differences (Control, MCI, AD) were highly significant by one-way ANOVA (*p* = 1.1 × 10⁻¹⁷). Samples with tTau/Aβ₁₋₄₂ ≥ 0.226 were classified as biomarker-positive (pink), whereas those with tTau/Aβ₁₋₄₂ < 0.226 were classified as biomarker-negative (yellow). **B)** Volcano plot showing differential abundance of 5,811 network proteins between biomarker-positive and biomarker-negative groups. The x-axis represents log₂ fold-change (biomarker-positive vs. biomarker-negative), and the y-axis shows −log₁₀ *p*-values. A total of 810 proteins were significantly increased and 763 decreased (*p* < 0.05) in biomarker-positive individuals, with points colored by plasma network module assignment. **C)** Biweight midcorrelation (bicor) volcano plot showing positive and negative associations between 5,811 plasma network proteins and AD biomarkers (CSF Aâ₁₋₄₂, CSF pTau181, CSF tTau/Aâ₁₋₄₂ ratio, and plasma pTau217). Correlations with CSF tTau are not shown due to space limitations. The x-axis represents bicor coefficients, and the y-axis shows −log₁₀ *p*-values. Significant proteins (*p* < 0.05) are colored by co-expression module. **D)** Heatmap showing 80 plasma proteins strongly correlated with AD biomarkers. The color scale represents Pearson correlation coefficients (green = positive; purple = negative), and circle size reflects significance. **E)** ROC curves comparing CSF biomarker-positive (*n* = 67) vs. biomarker-negative (*n* = 62) participants, showing AUCs for (a) a ten-protein panel (orange, AUC = 0.90), (b) plasma pTau217 alone (green, AUC = 0.91), and (c) the combination of pTau217 and the ten-protein panel (blue, AUC = 0.99). The combination significantly outperformed either alone by DeLong’s test (*p* = 0.004 and *p* = 0.0006). **F)** Bar plot showing mean SHAP values for each of the ten selected proteins; color boxes indicate each protein’s network module.

We next examined quantitative correlations between plasma proteins and AD biomarkers (CSF Aβ₁₋₄₂, tTau, pTau181, tTau/Aβ₁₋₄₂ ratio, and plasma pTau217). Although single-protein correlations were modest (peaking nearly |*cor*| ∼0.5), many associations were significant (*p* < 0.05) (**Figure 5C, Supplemental Table 25**). Positively correlated proteins were enriched especially in ECM, lipid metabolism, and keratin-related modules (M1, M2, M26), whereas negatively correlated proteins mapped to mitochondrial, proteostasis, and intracellular transport/cytoskeletal modules (M3, M4, M5, M6, M8, M11, M12). The 80 strongest correlating proteins are shown in **Figure 5D**.

Because individual proteins only partially captured CSF biomarker variation, we then evaluated whether a multivariate marker panel could improve classification. Using recursive feature elimination with a linear Support Vector Classifier (SVC), we identified a 10-protein panel that distinguished biomarker-positive from biomarker-negative individuals with an AUC of 0.90, comparable to plasma pTau217 alone (AUC = 0.91). Importantly, combining the panel with pTau217 significantly improved performance (AUC = 0.99; *p* = 0.004 vs. pTau217 alone), indicating that the panel provides complementary biological markers related to AD pathology (**Figure 5E–F**). These results show that the plasma proteome captures coordinated AD-related signatures that align with CSF biomarker status. Integrating proteomic markers with pTau217 further improves classification accuracy, indicating that plasma proteomics captures additional disease-relevant variation beyond pTau217 alone.

### Plasma Network Modules Are Preserved in the AD Brain and Show Concordant Disease-Related Changes

To determine whether plasma network modules capture biology that extends to the brain, we compared the 29 plasma modules with 44 previously defined consensus human brain proteomic modules (29). This comparison allowed us to assess whether groups of co-regulated proteins in plasma mirror coordinated molecular programs presenting in the CNS. Strikingly, plasma network showed substantially greater overlap with brain modules than previously reported using SomaLogic plasma data, where little to no preservation was observed (59). Here, 19 of 29 plasma modules significantly overlapped with one or more brain modules (**Figure 6A**). Many corresponding brain modules are highly correlated with AD pathology and cognitive decline and are enriched for astrocyte, microglial, neuronal, or endothelial markers (**Figure 6B-C**), suggesting that the plasma network, at least in part, reflects biological signals from brain cell types.

**Figure 6.**
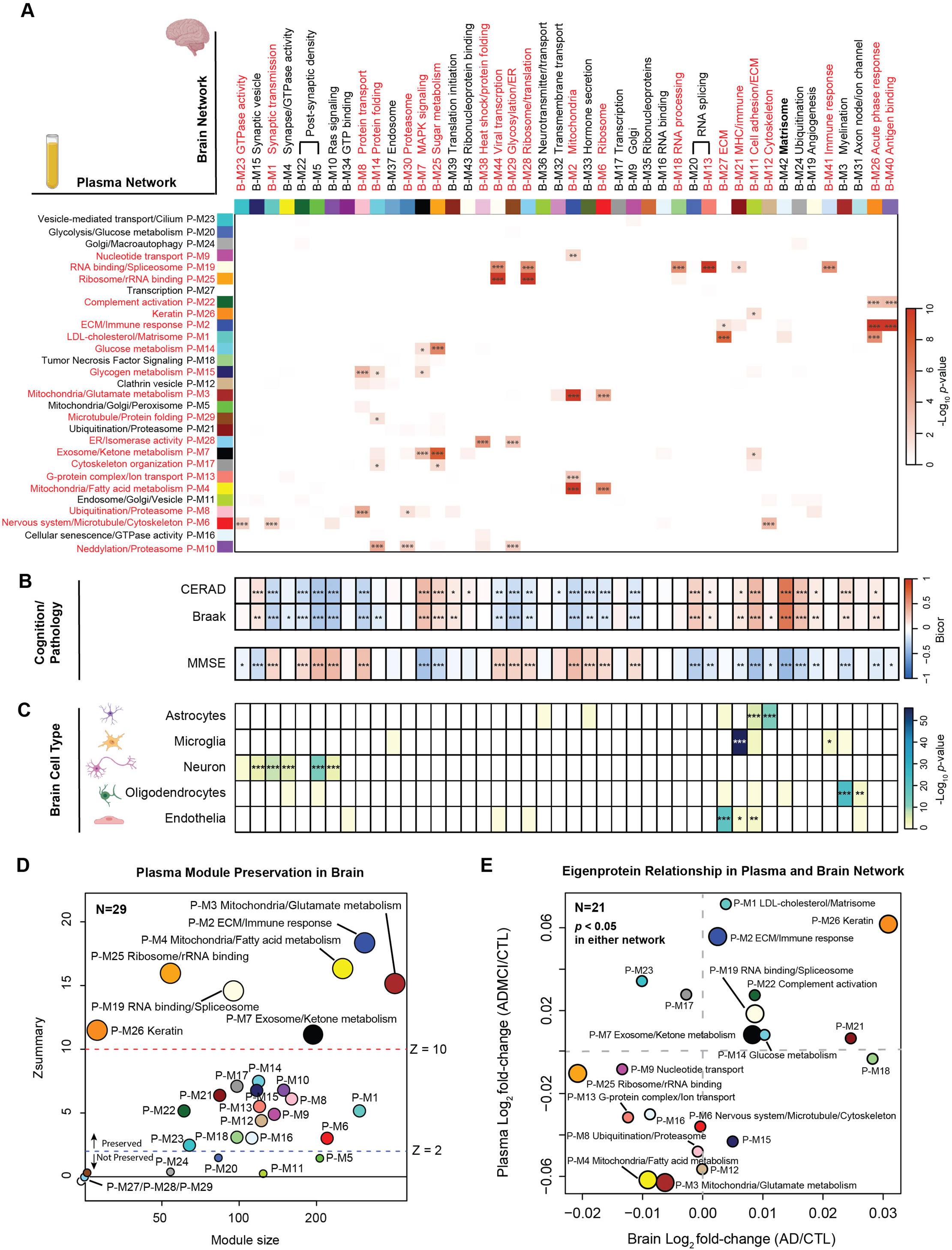
Concordance Between Plasma and Brain Protein Co-Expression Networks in AD. **A)** Overlap between 29 plasma co-expression network modules (y-axis) and 44 previously defined human brain consensus network modules (x-axis). The intensity of color shading represents the degree of protein overlap. Statistical significance was assessed by Fisher’s Exact Test (FET) followed by Benjamini–Hochberg FDR correction (\**p* < 0.05, \*\**p* < 0.01, \*\*\**p* < 0.001). Modules with significant overlaps are labeled red. **B)** The heatmap demonstrates the bicor of each brain module with CERAD, Braak, and MMSE cognitive scores (* *p* < 0.05, ** *p* < 0.01, *** *p* < 0.001). The M42 ‘Matrisome’ module exhibits the strongest correlation with AD pathology, and several synaptic modules (M1, M4, M5, and M22) display an overall decrease in AD brain. **C)** Cell-type enrichment of each brain module was assessed by FET using reference marker lists for astrocytes, microglia, neurons, oligodendrocytes, and endothelial cells. Color intensity reflects enrichment strength, and asterisks denote statistical significance (\**p* < 0.05, \*\**p* < 0.01, \*\*\**p* < 0.001). **D)** Preservation of plasma network modules within the brain proteome, quantified by Zsummary scores. Modules with Zsummary < 2 were considered not preserved (small circles, *n* = 7), 2 ≤ Zsummary < 10 were moderately preserved (medium circles, *n* = 15), and Zsummary ≥ 10 were highly preserved (large circles, *n* = 7). **E)** Relationship between module-level abundance changes in plasma and brain networks. For each module, log₂ fold-change (AD vs. CTL) in brain was plotted against log₂ fold-change (AD/MCI vs. CTL) in plasma. Only individuals classified as AD or MCI with positive CSF biomarkers and controls with negative CSF biomarkers were included. Among the 29 modules, 21 were significantly altered in AD (*p < 0.05*) in at least one dataset, with 18 showing concordant directional changes, indicating shared biological trends across compartments. Circle size corresponds to module preservation strength as in panel D.

We next examined whether the overall network architecture of plasma modules is preserved in brain. Using module preservation statistics from WGCNA (43), we found that roughly two-thirds of plasma modules (22 of 29) showed measurable preservation, and several were highly preserved (**Figure 6D**). This indicates that many plasma modules share underlying regulatory structure with their brain counterparts, suggesting partial coordination between the peripheral and central proteomes. However, preservation alone does not ensure concordant disease-associated alterations across compartments. We therefore compared AD-related module-level abundance changes in plasma and brain by examining their eigenprotein fold changes (biomarker-positive AD/MCI vs. biomarker-negative controls) for each module (**Supplemental Tables 26-27**). Among the 22 preserved modules, 21 were significantly altered in at least one dataset, and 18 showed concordant directional changes in both plasma and brain (**Figure 6E**). Modules increased in AD in plasma, including those linked to cholesterol metabolism, ECM/matrisome remodeling, lipid metabolism, and inflammation, were similarly elevated in AD brain. Conversely, modules decreased in AD in plasma, particularly those related to mitochondrial biology and proteostasis, were also reduced in AD brain. Together, these findings show that deep plasma proteomics capture protein co-expression patterns that are preserved in the brain and exhibit concordant disease-related changes across compartments.

### Plasma Proteome Partially Mirrors Brain Molecular Changes, Revealing Both Shared and Compartment-Specific Signatures in AD

After establishing module-level relationships between plasma and brain networks, we next prioritized shared module proteins to identify those showing the strongest concordant changes between compartments at the individual protein level. Using our previously reported deep brain proteomic analyses of 456 ROSMAP cases, which identified 8,956 proteins in brain (29), together with the 5,823 proteins quantified in plasma in the current study, we identified 4,887 (49.4%) proteins shared across both datasets (**Figure 7A, Supplemental Table 28**). Among these, 130 proteins (red) were significantly increased in both AD brain and plasma, whereas 293 (blue) were significantly decreased in both (**Figure 7B**). The remaining proteins showed compartment-specific patterns, with changes limited to one dataset or in opposite directions, indicating biological divergence between plasma and brain proteomes.

**Figure 7.**
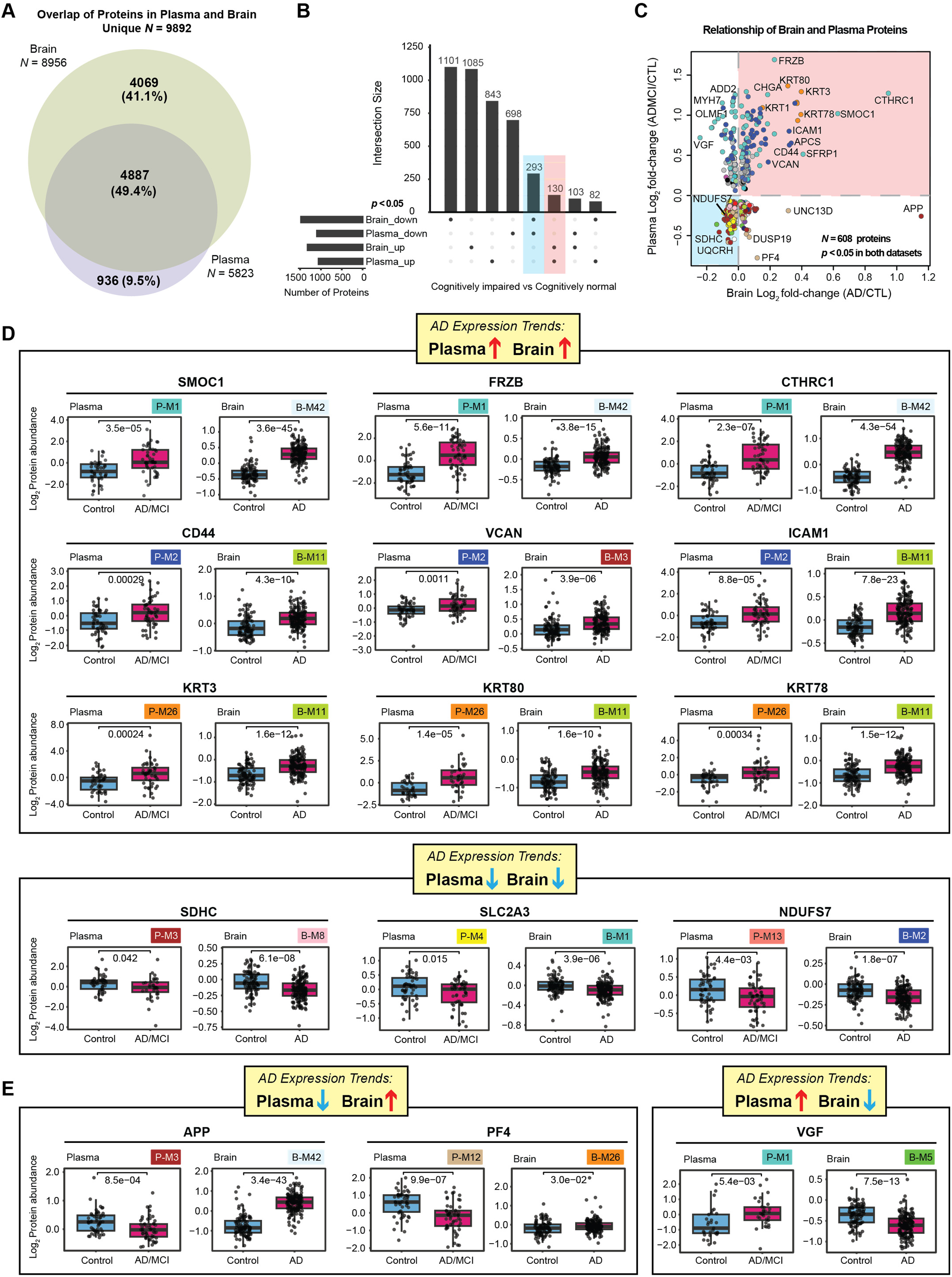
Shared and Divergent Proteomic Signatures in AD Plasma and Brain. **A)** Overlap of proteins quantified in brain (*N* = 8,956) and plasma (*N* = 5,823) datasets with less than 50% missing values. A total of 4,887 proteins (49.4%) were identified in both compartments. **B)** UpSet plot showing the number of proteins significantly increased or decreased in cognitively impaired versus cognitively normal individuals in brain and plasma. Cognitively normal cases were defined as control samples with CSF tTau/Aâ₁₋₄₂ < 0.226, while cognitively impaired cases included AD or MCI with CSF tTau/Aβ₁₋₄₂ ≥ 0.226. A total of 293 proteins were decreased in both plasma and brain (blue), and 130 proteins were increased in both (red). **C)** Scatter plot comparing protein log₂ fold-change in plasma (AD/MCI vs. CTL) and brain (AD vs. CTL). Only individuals classified as AD or MCI with positive CSF biomarkers and controls with negative CSF biomarkers were included. Proteins significantly altered in both datasets (*p* < 0.05, *N* = 608) are shown; 130 showed concordant increases and 293 exhibited decreases in both compartments. **D-E**) Representative examples of proteins with shared or opposing AD-related changes in plasma and brain. SMOC1, FRZB, CTHRC1, CD44, VCAN, ICAM1, KRT3, KRT80, and KRT78 increased in both compartments, whereas SDHC, SLC2A3, and NDUFS7 decreased. In contrast, APP, PF4, and VGF displayed opposite trends between plasma and brain. Statistical significance was determined by Student’s *t*-test for plasma and one-way ANOVA with Tukey post-hoc correction for brain.

To further assess individual protein-level correspondence, we compared log₂ fold-change in plasma and brain (biomarker-positive AD/MCI vs. biomarker-negative controls) for the 608 proteins significantly altered (*p* < 0.05) in both datasets (**Figure 7C, Supplemental Tables 29-30**). The resulting scatter plot revealed partial directional agreement, with a subset of proteins showing concordant changes in both compartments, while others showed opposite trends. This indicates that although some AD-related proteomic changes are shared between plasma and brain, others are compartment-specific or reflect differential regulation across the two biological systems. Notably, even proteins in the same network module did not always exhibit uniform directionality between plasma and brain, reinforcing the complexity of cross-compartment regulatory mechanisms. Representative examples of individual protein abundance changes in plasma and brain are shown in **Figure 7D-E**. Several matrisome and ECM-associated proteins, including SMOC1, FRZB, CTHRC1, and VCAN, were elevated in cognitively impaired individuals in both plasma and brain, consistent with extracellular matrix remodeling linked to amyloid and vascular pathology (14, 41, 60). Immune-related proteins such as ICAM1 and CD44 showed similar concordant increases across compartments, potentially reflecting shared endothelial and glial inflammatory responses (61–63). Interestingly, multiple keratin family proteins (KRT3, KRT80, KRT78) also displayed increased abundance in both plasma and brain, potentially linked to a systemic epithelial or stress-related response in AD (64–66). In contrast, mitochondrial and metabolic proteins such as SDHC, SLC2A3, and NDUFS7 were consistently reduced, consistent with a hypometabolic phenotype in AD (67–70). Together, these patterns show that key pathways identified by co-expression analysis, such as ECM remodeling, immune response, and mitochondrial dysfunction, are reflected in overlapping protein subsets across plasma and brain. However, some proteins (e.g., APP, PF4, VGF) exhibit opposite directions of change, highlighting complex central–peripheral dynamics. Overall, plasma and brain share broad AD-related signatures at the pathway level, while individual proteins vary in direction and magnitude, indicating both concordant signals and compartment-specific regulation.

## Discussion

In this study, we applied a microbead enrichment–based DIA-MS approach to deeply profile plasma samples from over 200 individuals spanning the AD clinical spectrum. Importantly, paired CSF samples were available for a substantial subset of participants, allowing stratification not only by clinical diagnosis but also by CSF amyloid and tau biomarker status. Using a network-based approach, we identified groups of co-expressed proteins that reflect key biological processes associated with AD and demographic factors. These plasma protein patterns closely aligned with CSF and plasma biomarkers of AD. Using machine learning approaches, we nominated a ten-protein plasma panel that distinguishes CSF biomarker-positive from -negative individuals and significantly improves the classification performance of plasma pTau217 alone. Finally, integration with human brain proteomic networks revealed substantial overlap between peripheral and central molecular changes, demonstrating that the plasma proteome reflects core features of brain pathology while also capturing broader systemic responses to AD.

A major strength of this study lies in the analytical depth, reproducibility and consistency achieved through the combination of standardized microbead enrichment-based sample processing and DIA-MS data acquisition. We reliably measured 5,823 proteins, comparable to the SomaScan platform (41) and spanning 10 orders of magnitude in plasma abundance, reaching concentrations as low as 2.90 pg/ml, including low-abundance proteins like SYNJ1 and PLCB4. Cross-platform comparison showed positive correlations between overlapping proteins in our method compared to the SomaScan (*cor* = 0.7) or Heparin TMT-MS (*cor* = 0.78) platform. Technical and instrumental variance analyses also confirmed high consistency of the data, supporting the reliability of our downstream network and biomarker analyses. The use of CSF and plasma biomarker data within the same cohort provided a unique opportunity to directly link peripheral proteomic changes to the established measures of AD pathology.

Plasma protein network analysis identified biologically coherent modules capturing diverse aspects of AD pathology and demographic variation. In total, 29 co-expression modules were defined, representing key processes including extracellular matrix remodeling, immune response, mitochondrial metabolism, vesicle trafficking, and cytoskeletal organization. Modules such as M1 (LDL-cholesterol/matrisome), M2 (ECM/immune response), and M26 (keratin) showed the strongest positive associations with AD diagnosis, APOE4 gene dose, and CSF or plasma biomarkers, whereas mitochondrial and proteostasis-related modules (M3, M4, M5, M6, M8, M11, M12) decreased in AD plasma. When comparing the plasma network with a consensus AD brain proteomic network (15, 29), approximately two-thirds of plasma modules were preserved in the brain network, suggesting conserved co-expression topology across the two compartments. Importantly, many of these preserved modules were enriched for protein signatures characteristic of brain cell types, including astrocytic, microglial, endothelial, and neuronal proteins, implying that a subset of plasma proteins may reflect brain-derived or CNS-related processes. These relationships likely arise from extracellular vesicles, leakage across the blood–brain barrier, or peripheral responses to central pathology.

One key observation in our study was the consistent decrease of mitochondrial-related modules across both plasma and brain proteomes. Mitochondrial dysfunction is a well-established feature in AD pathology (71–76), with extensive evidence implicating impairments in oxidative phosphorylation, lowered respiration rates, and altered electron transport chain (ETC) activity linked to neuronal injury and synaptic degeneration (45, 77, 78). Although plasma is traditionally considered cell-free, recent studies have reported alterations in circulating mitochondrial markers, including changes in cell-free mitochondrial DNA levels (79–82), as well as mitochondrial-related pathway perturbations detectable in plasma and CSF (53, 83), suggesting that aspects of mitochondrial dysfunction in AD can be monitored peripherally. Notably, the plasma mitochondrial module substantially overlapped with the brain mitochondrial module and exhibited a concordant decrease in AD. This suggests that the plasma mitochondrial signal does not simply reflect leakage of brain-derived proteins into circulation, but rather represents conserved, cross-tissue changes that are coordinately disrupted in AD. Given that mitochondrial pathways are broadly expressed across cell types and organs, their shared reduction likely reflects systemic bioenergetic stress rather than cell-type–specific neuronal loss alone. Our findings therefore support the concept that systemic mitochondrial alterations mirror central neurodegenerative processes while also reflecting broader metabolic dysfunction. Moreover, it raises the possibility of developing accessible, blood-based biomarkers that reflect CNS metabolic dysfunction, which could be a promising way for early detection and disease monitoring in AD.

Keratin module (M26) is another interesting case that exhibited significant increase in both AD plasma and brain. This module was dominated by epithelial cytoskeletal proteins, including KRT1, KRT9, KRT78, and KRT80. Notably, keratin 9 (KRT9) has been previously detected in both plasma and CSF of AD patients and was included in a CSF protein panel to discriminate AD patients from controls (84–86). It was suggested that dysregulation of KRT9 in blood and CSF may be linked to blood–brain barrier (BBB) disruption and proteostasis changes in AD pathology, and that keratin proteins interact with other AD-related pathways at the molecular network level, even though their biological role in AD remains to be fully clarified (84, 87). Although keratins are often regarded as epithelial proteins and are usually considered potential contaminants in proteomic studies, transcriptomic analyses indicate that multiple keratin genes are expressed across diverse brain cell types, including neurons, astrocytes, microglia, oligodendrocytes, and endothelial cells (44). Consistent with this, the plasma keratin module overlapped with a brain “cell adhesion/ECM” module enriched for endothelial markers, including keratin proteins. This suggests that increased keratin abundance may reflect neurovascular remodeling and endothelial activation rather than peripheral epithelial contamination. Endothelial dysfunction and BBB breakdown are increasingly recognized as key components of AD pathogenesis, contributing to impaired amyloid clearance, inflammatory cell trafficking, and altered molecular exchange between the CNS and circulation (88–92). The enrichment of endothelial signatures supports a vascular contribution to the plasma keratin signal. Our findings recapitulate prior observations of KRT9 and extend them by demonstrating a coordinated increase across multiple keratin family members.

Interestingly, directional agreement between plasma and brain was only partial at the individual protein level. While some proteins showed concordant changes across compartments, others exhibited opposite or compartment-specific patterns, reflecting complex CNS–peripheral dynamics. For example, VGF decreased in AD brain but increased in plasma in this study, consistent with neuronal loss in the brain and potential compensatory secretion or altered trafficking in the periphery (93, 94). These differences may also reflect systemic responses to neurodegeneration, including immune, metabolic, and vascular changes. Rather than weakening the disease relevance of plasma signals, this divergence highlights the interplay between central and peripheral biology, supporting the view of AD as a multi-system disease (61, 67, 95–97).

Proteins within these network modules showed similar directional changes when stratifying samples by CSF biomarker positivity, further supporting their disease relevance. Plasma pTau217, which has emerged as a robust and scalable biomarker for amyloid pathology (8, 98, 99), served as a critical anchor for implying plasma-brain correspondence. We showed that many proteins associated with pTau217 were also correlated with CSF tTau/Aβ₁₋₄₂ ratio, confirming that diverse plasma proteomic signatures align closely with traditional AD biomarker profiles. Beyond single-protein associations, we identified a ten-protein panel that demonstrated comparable classification performance to plasma pTau217 alone. Notably, when combined with pTau217, this panel significantly enhanced the discrimination of CSF biomarker-positive versus biomarker-negative individuals, underscoring the added value of multiplexed plasma proteomic signatures in improving diagnostic accuracy and stratification in Alzheimer’s disease.

Finally, this study has several limitations. Although we achieved deep plasma proteome coverage, the cross-sectional design precludes determination of whether these changes precede or follow disease progression. Longitudinal studies and replication in independent cohorts will be required to establish predictive value and generalizability. In addition, while plasma signatures align with AD-related brain changes, we cannot infer tissue of origin given the heterogeneous sources of circulating proteins; approaches such as extracellular vesicle-based or cell-type–resolved proteomics may help address this limitation. The cohort size and composition may also limit power to detect subtle effects and to fully resolve interactions with sex and race. Furthermore, technical factors, including microbead enrichment bias and platform-specific detection differences, may influence protein representation and cross-platform comparability. Finally, while network and biomarker associations are robust, they remain correlative and do not establish causal mechanisms linking plasma proteins to AD pathobiology.

## Conclusion

In conclusion, this study provides a system-level characterization of the plasma proteome in AD and its relationship to brain pathology. By defining reproducible network modules and identifying proteins and pathways that track with central AD processes, we link peripheral and brain proteomic changes. These findings support microbead-enriched MS-based plasma proteomics as a scalable approach for biomarker discovery and biological insight, enabling more accessible and integrative strategies to detect and monitor AD.

## Supplemental Figure Legends

**Supplemental Figure 1.**
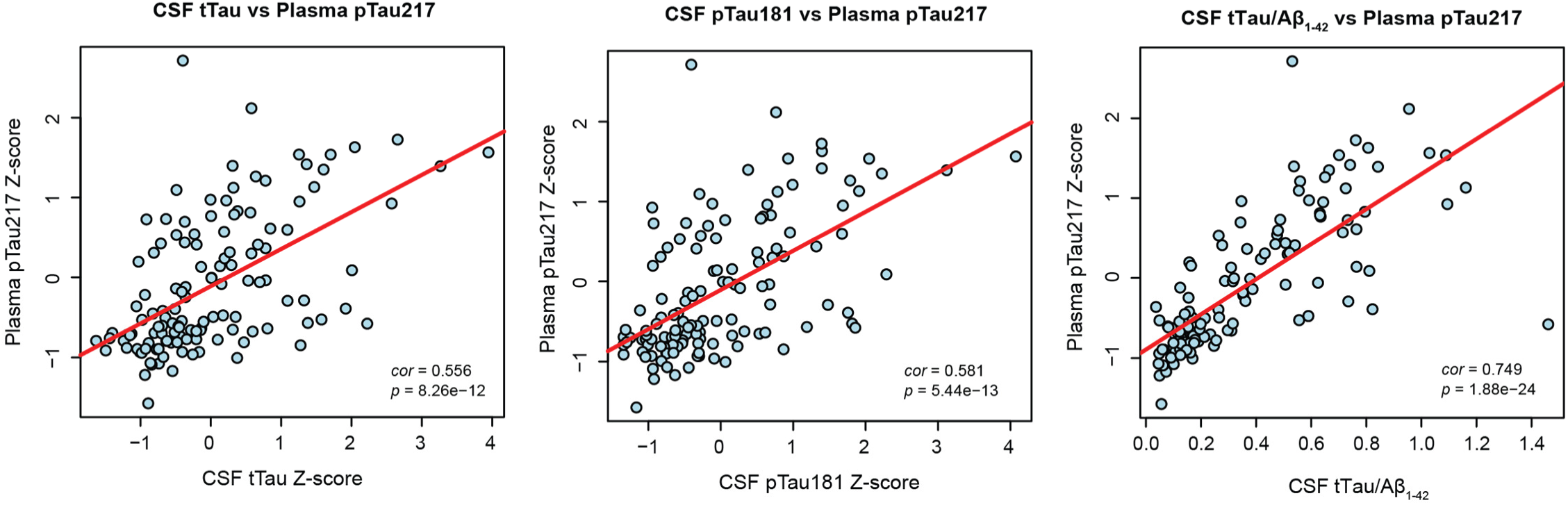
Correlations Between Plasma pTau217 and CSF AD Biomarkers. Scatter plots showing Pearson correlations between Z-scored plasma pTau217 and CSF AD biomarkers: CSF total tau (tTau; *r* = 0.56, *p* = 8.3 × 10⁻¹²), CSF phospho-tau181 (pTau181; *r* = 0.58, *p* = 5.4 × 10⁻¹³), and the CSF tTau/Aβ₁₋₄₂ ratio (*r* = 0.75, *p* = 1.9 × 10⁻²⁴). Statistical significance of correlations was assessed using Student’s *t*-test.

**Supplemental Figure 2.**
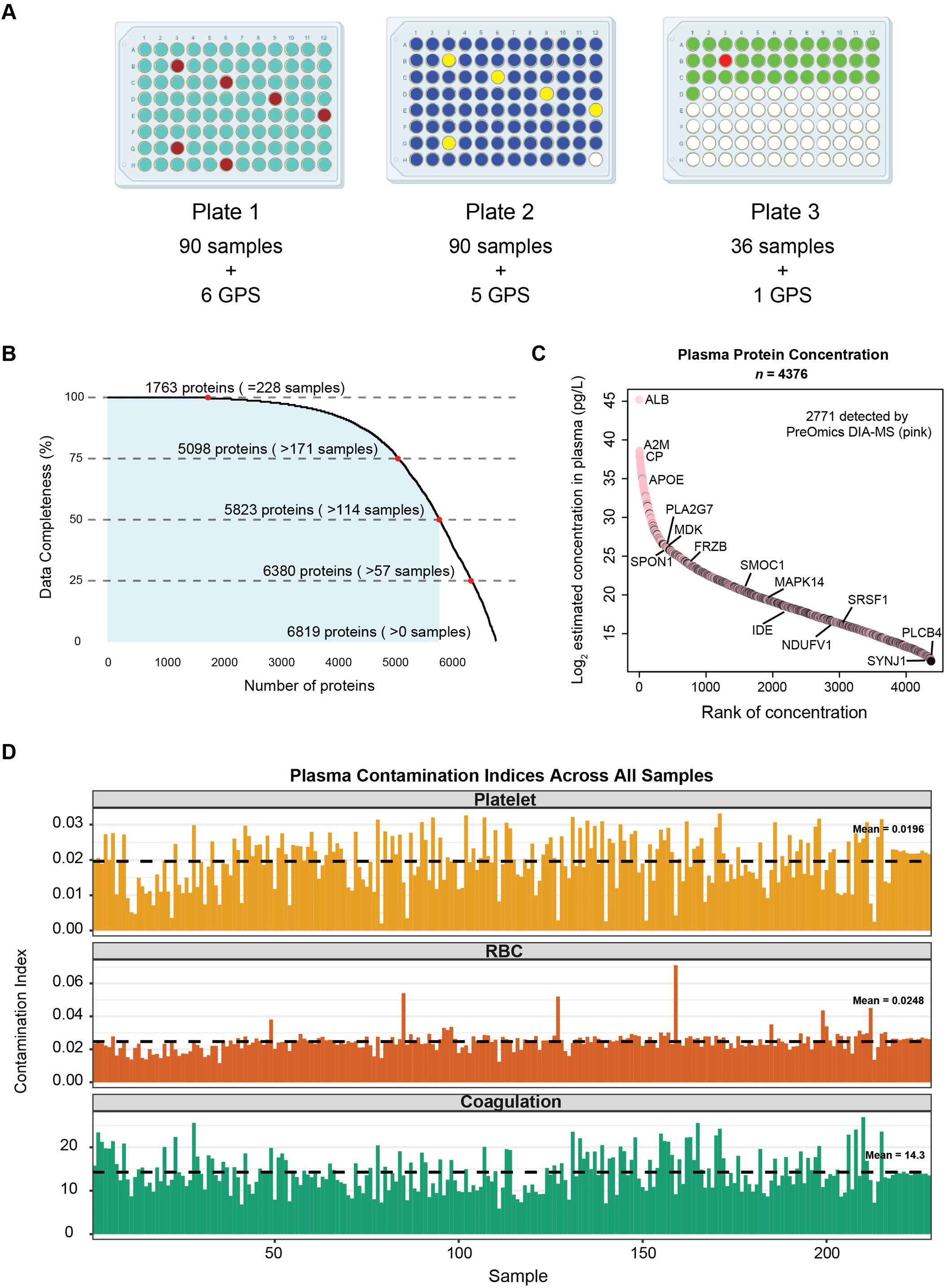
Overview of Data Depth and Sample Quality. **A)** Distribution of plasma samples (*n* = 216, including 2 duplicates) and Global Pooled Standards (GPS) samples (*n* = 12) across three 96-well plates. Plate 1 includes 90 samples (turquoise) and 6 GPS (brown); Plate 2 includes 90 samples (blue) and 5 GPS (yellow); Plate 3 includes 36 samples (green) and 1 GPS (red). **B)** Protein measurement completeness across 228 samples: 1,763 proteins were quantified in all 228 samples (no missing values, i.e., 100% completeness); 5,823 proteins were detected in more than half of the samples and were retained for downstream analysis. **C)** Of the 4,376 plasma proteins with estimated concentrations reported in the Human Protein Atlas database (47), 2,771 were detected using our PreOmics DIA-MS workflow. All 4,376 proteins were ranked by their estimated concentrations (log2-transformed, pg/L), with proteins detected by DIA-MS highlighted in pink and those not detected shown in black. Protein abundances span approximately ten orders of magnitude, and low-abundance proteins such as SYNJ1 and PLCB4 were successfully detected. **D)** Potential contamination from platelets, erythrocytes (RBC), and coagulation-related proteins was evaluated using Baize software on raw, unnormalized protein abundance data from all 228 plasma samples, including technical duplicates. Each bar represents one sample, and dashed horizontal lines indicate the cohort mean contamination index for each category.

**Supplemental Figure 3.**
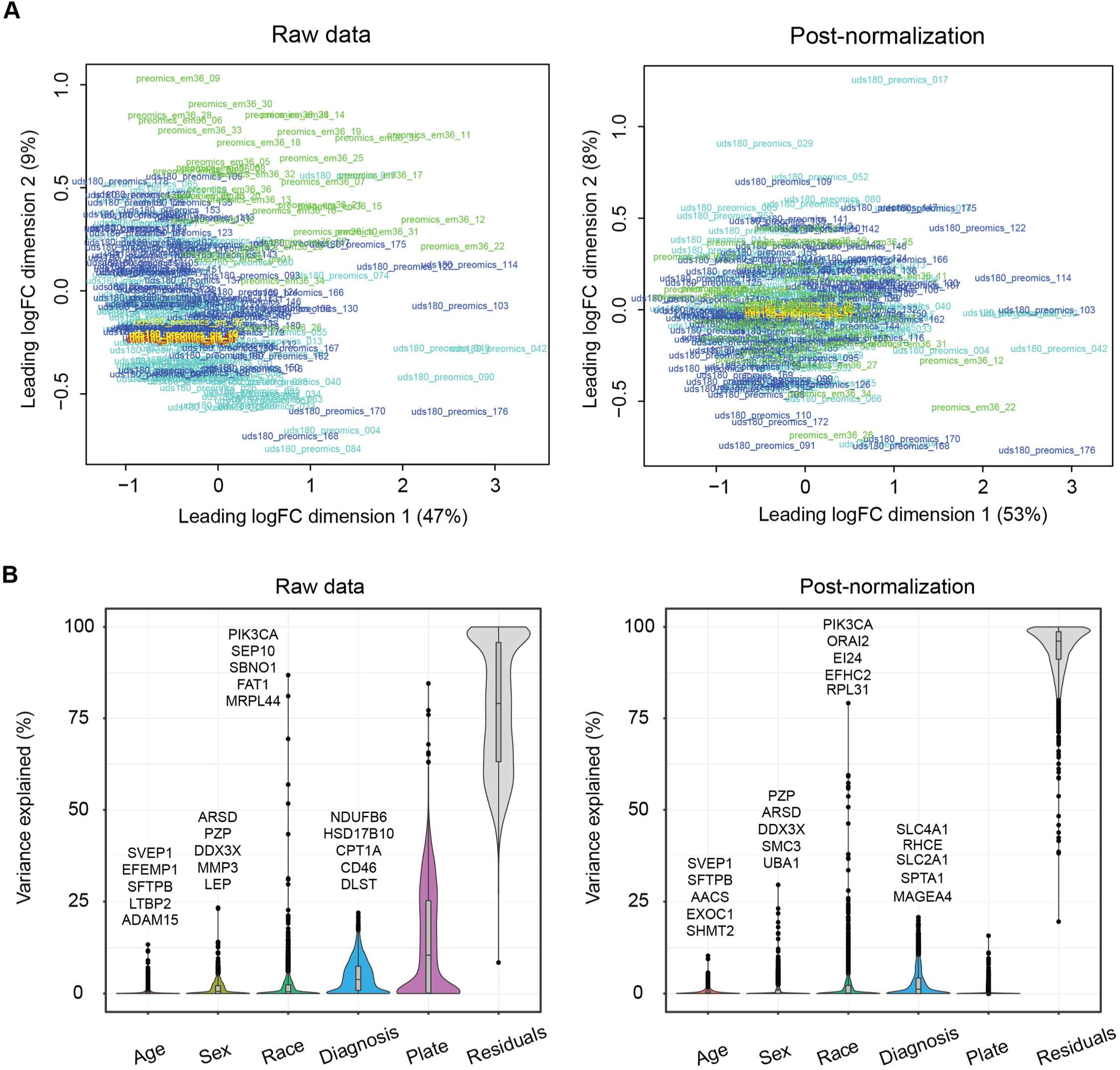
TAMPOR Normalization Effectively Removes Plate-Related Variance While Preserving Biological Signals in Plasma DIA-MS Data. **A)** Protein abundance data from 216 plasma samples and 12 global pooled standards (GPS) were normalized using the Tunable Approach for Median Polish of Ratios (TAMPOR). Multidimensional scaling (MDS) plots display sample clustering before (left) and after (right) normalization. In the raw data, samples cluster by plate—Plate 1 (blue), Plate 2 (turquoise), Plate 3 (green), and GPS samples (brown, yellow, or red)—indicating strong plate effects. After TAMPOR normalization, samples cluster tightly regardless of plate, demonstrating effective removal of plate-related variance. **B)** Variance partition analysis before (left) and after (right) TAMPOR normalization. The y-axis shows the percentage of variance explained by each factor (age, sex, race, diagnosis, plate, and residuals). After normalization, variance attributed to plate is markedly reduced, while biological variance is preserved. The top five proteins most influenced by age, sex, race, and diagnosis are labeled.

**Supplemental Figure 4.**
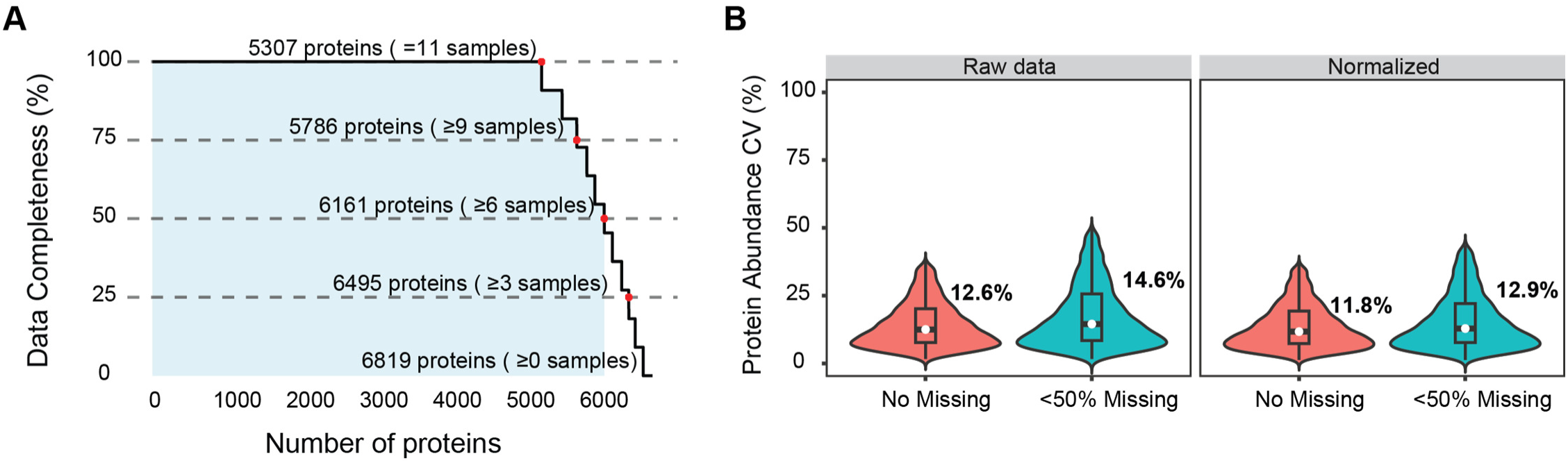
High Technical Reproducibility and Low Post-Normalization Variance across Global Pooled Standard (GPS) Samples. **A)** Data completeness across 11 GPS samples. A total of 5,307 proteins were quantified in all GPS samples (no missing values), and 6,161 proteins were detected in more than half of the samples (<50% missing). **B)** Violin plots showing protein abundance variability (coefficient of variation, CV) across GPS before and after normalization (instrument variance). Proteins quantified without missing values (orange) and those detected in at least 6 of the 11 GPS (green) are shown. Median CVs are labeled for each plot. For visualization purposes, CV values outside the range of mean ± 2 standard deviations (∼5% for each group) are not shown in the violin plots. All statistics are calculated using the full dataset, including all proteins.

**Supplemental Figure 5.**
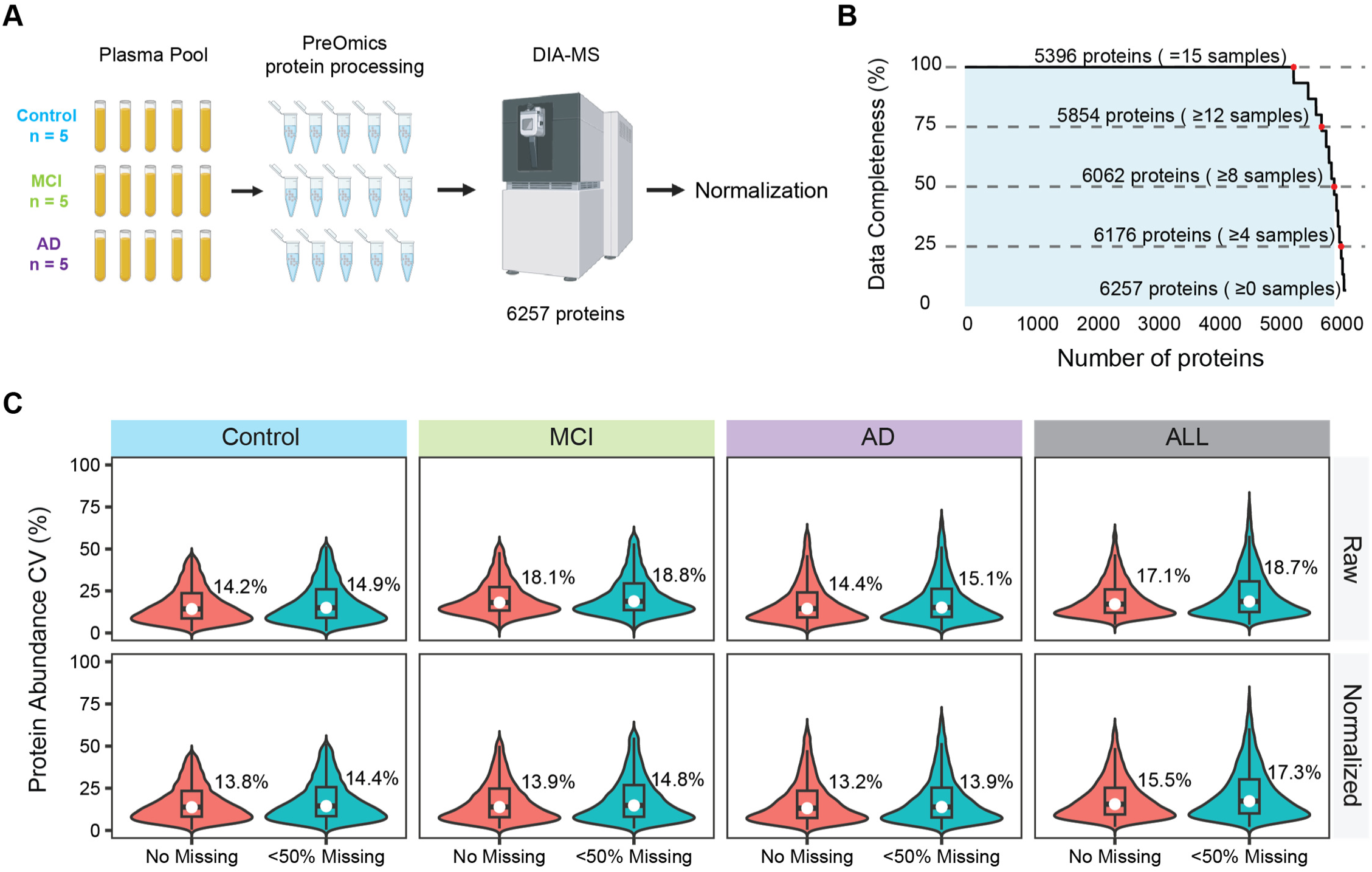
Microbead Enrichment Workflow Demonstrates High Proteome Coverage and Reproducibility. **A)** Workflow of 15 pooled plasma samples (5 Control, 5 MCI, 5 AD) processed the same way as cohort samples using the PreOmics DIA-MS pipeline, identifying 6,257 proteins. **B)** Data completeness across the 15 pooled plasma samples, with 5,396 proteins quantified in all samples (no missing values) and 6,062 detected in more than half of the samples. **C)** Violin plots showing distributions of protein abundance variability (coefficient of variation, CV) before and after normalization across pooled plasma samples (technical variance). Proteins quantified without missing values are shown in orange, and those detected in at least 8 of the 15 pooled plasma samples are shown in green. Median CVs are labeled for each plot. For visualization purposes, CV values outside the range of mean ± 2 standard deviations (∼1%-5% for each group) are not shown in the violin plots. All statistics are calculated using the full dataset, including all proteins.

**Supplemental Figure 6.**
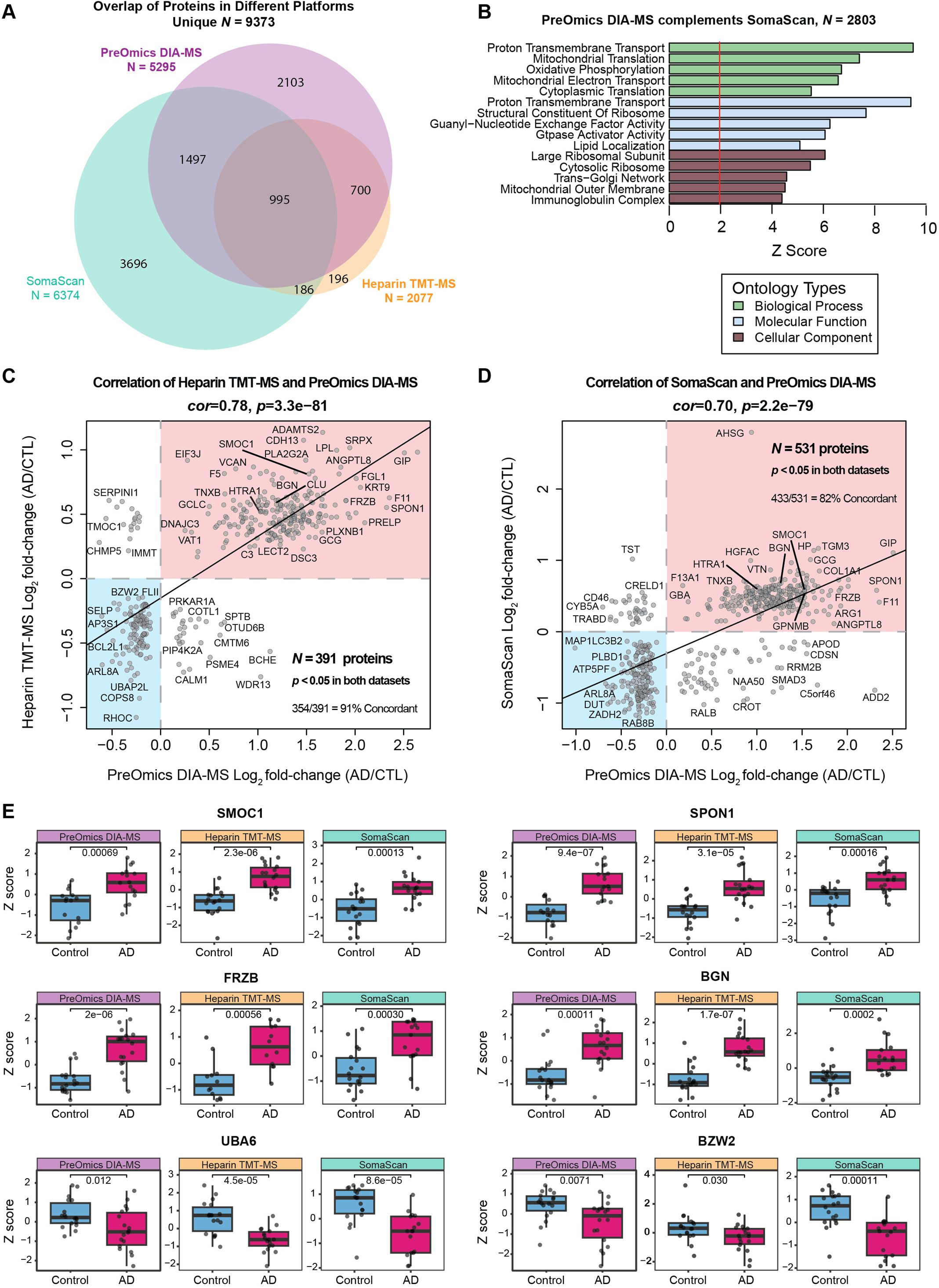
Cross-Platform Validation of AD Plasma Proteomics Across Mass Spectrometry and Aptamer-Based Assays. **A)** Overlap of plasma proteins quantified across three independent platforms using overlapping samples: microbead-enriched DIA-MS (PreOmics DIA-MS, *N* = 5,295 proteins; Control = 18, AD = 18), heparin-enriched TMT-MS (Heparin TMT-MS, *N* = 2,077 proteins; Control = 18, AD = 18), and aptamer-based assays (SomaScan, *N* = 6,374 proteins; Control = 18, AD = 17). Only proteins detected in at least 18 samples per platform were included. **B)** Gene Ontology (GO) enrichment of 2,803 proteins uniquely identified by PreOmics DIA-MS relative to SomaScan. These proteins were enriched for mitochondrial and translational processes, including mitochondrial translation and mitochondrial outer membrane, highlighting complementary coverage of intracellular biology beyond aptamer-based assays. **C–D)** Pairwise correlations (Pearson correlation) of log₂ fold-change (AD vs. Control) between PreOmics DIA-MS and Heparin TMT-MS (**C**, *r* = 0.78, *p* = 3.3 × 10^-81^; *N* = 391 proteins, 91% concordant) and between PreOmics DIA-MS and SomaScan (**D**, *r* = 0.70, *p* = 2.2 × 10^-79^; *N* = 531 proteins, 82% concordant). Only proteins overlapping and significantly altered (*p* < 0.05) in both datasets were included. **E)** Boxplots showing consistent protein abundance changes in AD across all three platforms for SMOC1, SPON1, FRZB, BGN, UBA6, and BZW2. Statistical significance was determined using Student’s *t*-test.

